# Importance of confirmatory test characteristics in optimizing community-based screening for tuberculosis: An epidemiological modeling analysis

**DOI:** 10.1101/2025.05.09.25327330

**Authors:** Lukas E Brümmer, Theresa S. Ryckman, Sourya Shrestha, Florian M Marx, William Worodria, Devasahayam J Christopher, Grant Theron, Adithya Cattamanchi, Claudia M Denkinger, David W Dowdy, Emily A Kendall

## Abstract

**Background:** Current active case-finding (ACF) efforts for tuberculosis (TB) are limited by the costs, operational barriers, and sensitivity of available tools to confirm a TB diagnosis. However, it is not well understood which of these limitations has the greatest epidemiological relevance and might therefore warrant prioritization in test development.

**Methods:** We developed a state-transition model of a one-time, community-based ACF intervention, with a fixed budget of one million United States dollars for screening and confirmatory testing. Assuming an adult population with four time the national prevalence of Uganda, we compared the impact of this intervention on TB diagnoses, mortality, and transmission when using a currently available confirmatory test (mirroring sputum-based Xpert Ultra) versus an improved confirmatory test. We considered the following test improvements: (1) increased sensitivity (from 69% to 80%), (2) non-sputum specimen type (increasing specimen availability from 93% to 100%), (3) immediate turn-around of test results (increasing delivery of positive results from 91% to 100%), (4) reduced costs (from $20 to $9 per confirmatory test). For those individuals not included in ACF efforts, TB outcomes under routine care were informed by recent natural history models.

**Results:** In a simulated target population of 400,000 adults, 6,421 (1.6%; 95% uncertainty range [UR] 5,316-7,531) had TB disease, and 873 (612-1,182) were projected to die of TB in the absence of ACF. Assuming current tests, ACF efforts could reach 83,808 (59,388-118,601; 21% of the target population) people under the allotted budget, connecting 651 (429-983) individuals with TB to treatment and averting 76 (39-132) deaths. Of all hypothetical confirmatory test improvements modeled, higher diagnostic sensitivity most increased the number of people with TB who received treatment as a result of ACF (by 14% [4-26%]). However, considering mortality or transmission as a metric, the largest reductions resulted from tests that provided immediate turn-around of results (by 11% [5-18%]).

**Conclusion:** Making confirmatory tests for community-based TB screening more accessible and rapid may lead to greater population health benefits than further increasing sensitivity. Nonetheless, achieving large (>20%) increases in the health impact of ACF will require improvements to components of ACF other than the confirmatory diagnostic test.

## INTRODUCTION

Every year, an estimated four million people with new tuberculosis (TB) are not notified to public health authorities and many of these people are never diagnosed or treated [1]. Community-based screening or active case-finding (ACF) can potentially reduce TB mortality and transmission potential, by detecting and linking people with TB to treatment that would have otherwise been missed or only diagnosed after they have transmitted TB to others [2]. The uptake and epidemiological impact of ACF, however, is limited by deficiencies of currently available diagnostic tests [3].

Most ACF algorithms begin with a low-cost screening test or symptom survey, and thus require a second, more specific test to confirm positive screening results [4]. Currently, advanced tools for confirmatory testing include molecular sputum tests such as Xpert Ultra MTB / RIF (Cepheid, Sunnyvale, CA, United States; “Xpert Ultra”) [5–7], but even the best tools available are subject to several shortcomings. First, high testing costs [8] may limit the number of people who can be tested under a given budget. In addition, the need for a sputum specimen hampers testing reach and the long turn-around times associated with off-site and high-volume testing can inhibit people from receiving their test results [6].

Furthermore, even molecular tests designed for high sensitivity leave some individuals with low bacillary load undetected [9]. While many of the TB diagnostics development efforts that are underway could lead to tools that would overcome these shortcomings, it is not clear which test characteristics could provide the largest benefit when used for the confirmatory step in ACF algorithms – and might therefore warrant higher prioritization during test development.

To guide researchers and public health decision makers in developing valuable diagnostics for ACF, we developed a model exploring the importance of confirmatory test characteristics in the context of community-based screening for TB. We consider an illustrative community-based ACF intervention that begins with chest x-ray screening in a high-TB-risk population in a setting similar to Uganda. We then estimate the impact of ACF on TB mortality, transmission potential and treatment initiation, when using Xpert Ultra versus various enhanced, hypothetical assays as the confirmatory test.

## METHODS

### Model design and TB disease course

We construct a model of an ACF intervention for adults, conducted on a background of routine TB care, to estimate the epidemiological impact of improving different characteristics of the confirmatory test used in ACF (Table 1). The results of ACF (detected and linked to treatment, or not) are estimated using a compartmental model (Figure 1A), which we place in a larger decision tree to project the individual-level clinical outcomes as well as the cumulative duration of TB disease under routine care and with the addition of ACF (Figure 1B). We differentiate prevalent TB by three characteristics: (i) high- or low-bacillary load (corresponding to positive or negative smear-microscopy status), (ii) presence of chronic cough (“cough positive” if a person with TB would report cough for more than two weeks, or “cough negative” otherwise), and (iii) HIV infection status (“HIV positive” or “HIV negative”). We assume that people with high bacillary loads are both more (i.e., fourfold [10]) infectious and more likely to test positive with any given confirmatory test [4, 9]. As an illustrative setting, we consider a hypothetical high-prevalence population of 400,000 adults in Uganda who are targeted for screening. They are modeled as having a TB prevalence of 1.6%, four times the national average [11], and a joint distribution of TB bacillary load, HIV status, and chronic cough reflecting nationwide patterns [11–14] (Table 2).

**Figure 1.**
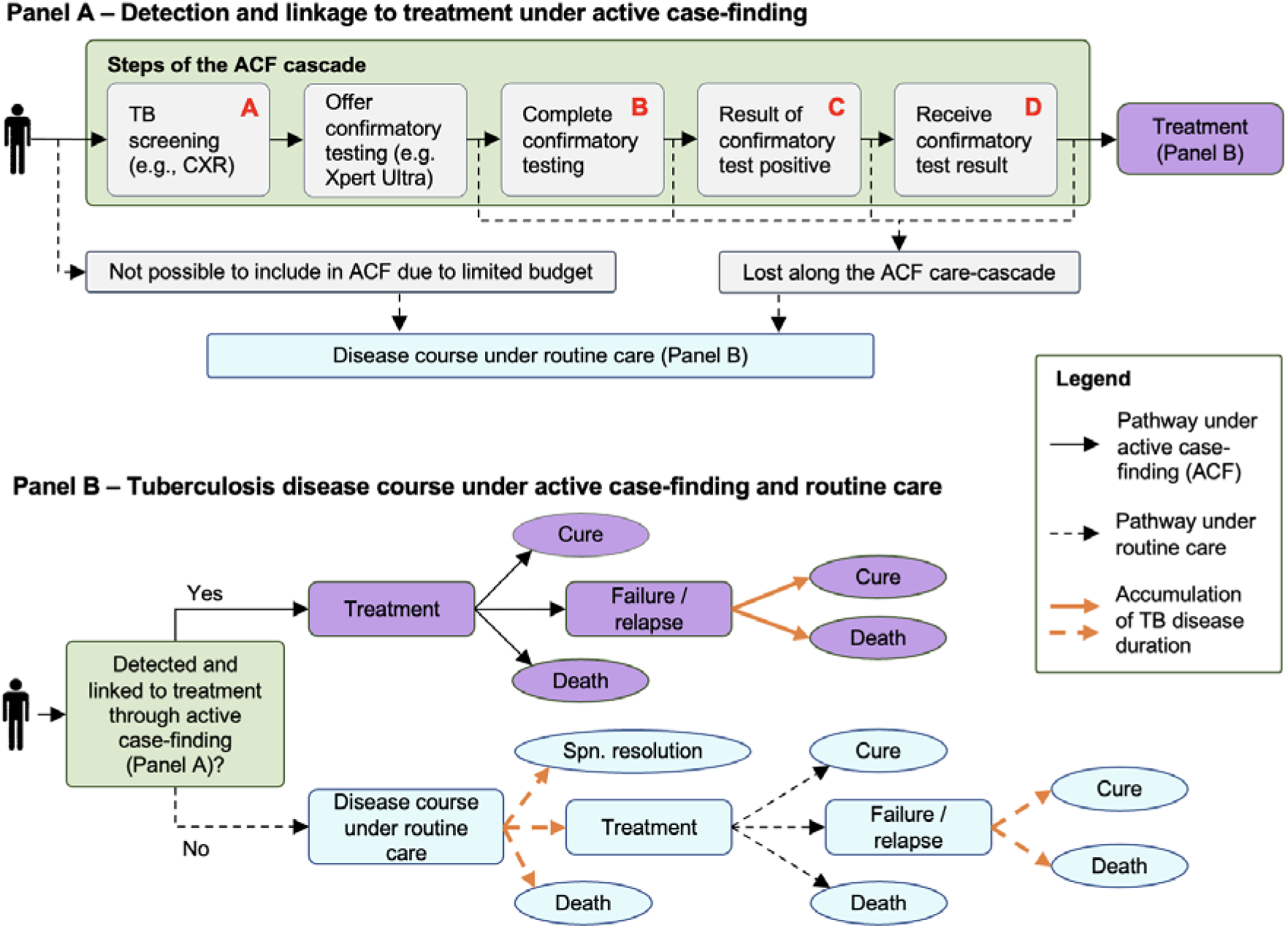
Model structure. We model a one-time active case finding (ACF) intervention, consisting of one-time screening (modeled as chest X-ray) plus confirmatory testing for those who screen positive (Panel A). We consider four different improvements to the confirmatory test that could increase the number of people successfully diagnosed and treated under ACF efforts: (A) reduced testing costs from $20 to $10 per test; (B) a non-sputum specimen type, increasing confirmatory specimen availability from 93% to 100%; (C) a point-of-care format, increasing receipt of confirmatory test results from 91% to 100%; (D) increased test sensitivity from 69% to 80%. Improvement A increases the number of people that can be included in the ACF efforts under a budget constraint, while improvements B, C, and D directly enhance the confirmatory testing process (Table 1). We estimate TB disease outcomes under ACF or – for people with TB who are not screened or not detected through ACF – under routine care using a decision-tree model, projecting TB to eventually end in cure, spontaneous resolution, or death (Panel B). In addition, we project smear-negative and smear-positive disease duration until any of these endpoints are reached (depicted through the bold, orange arrows in Panel B).

**Table 1.**
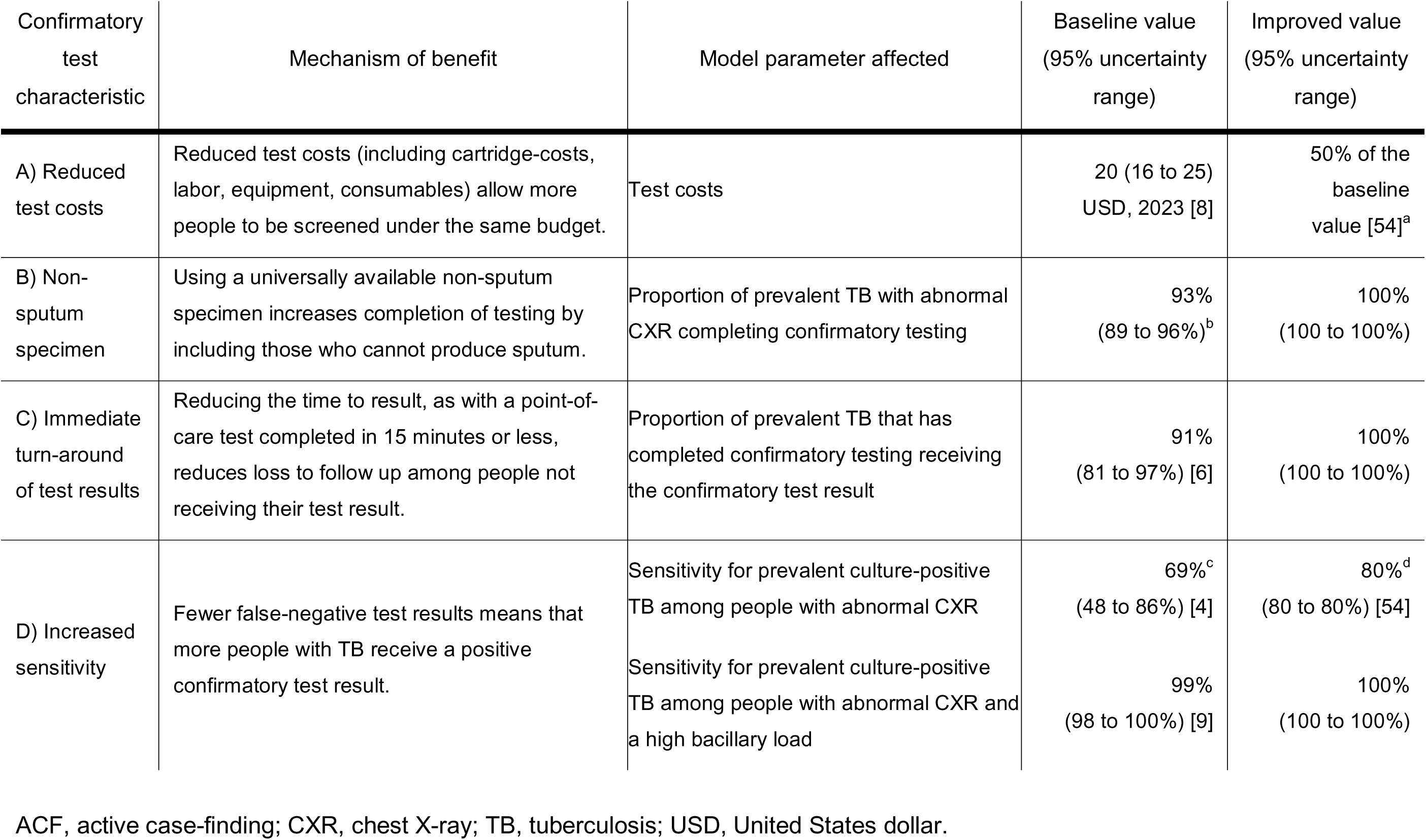
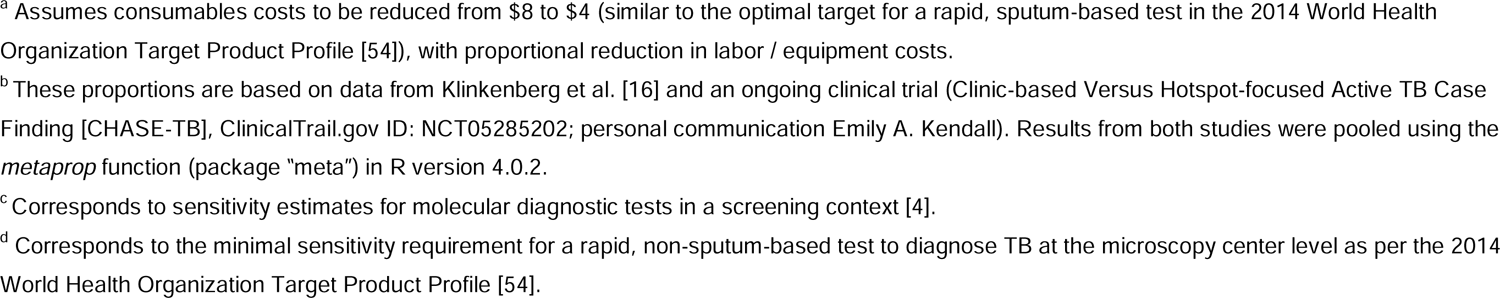
Hypothetical improvements of a test to confirm TB during active case-finding efforts.

**Table 2.**
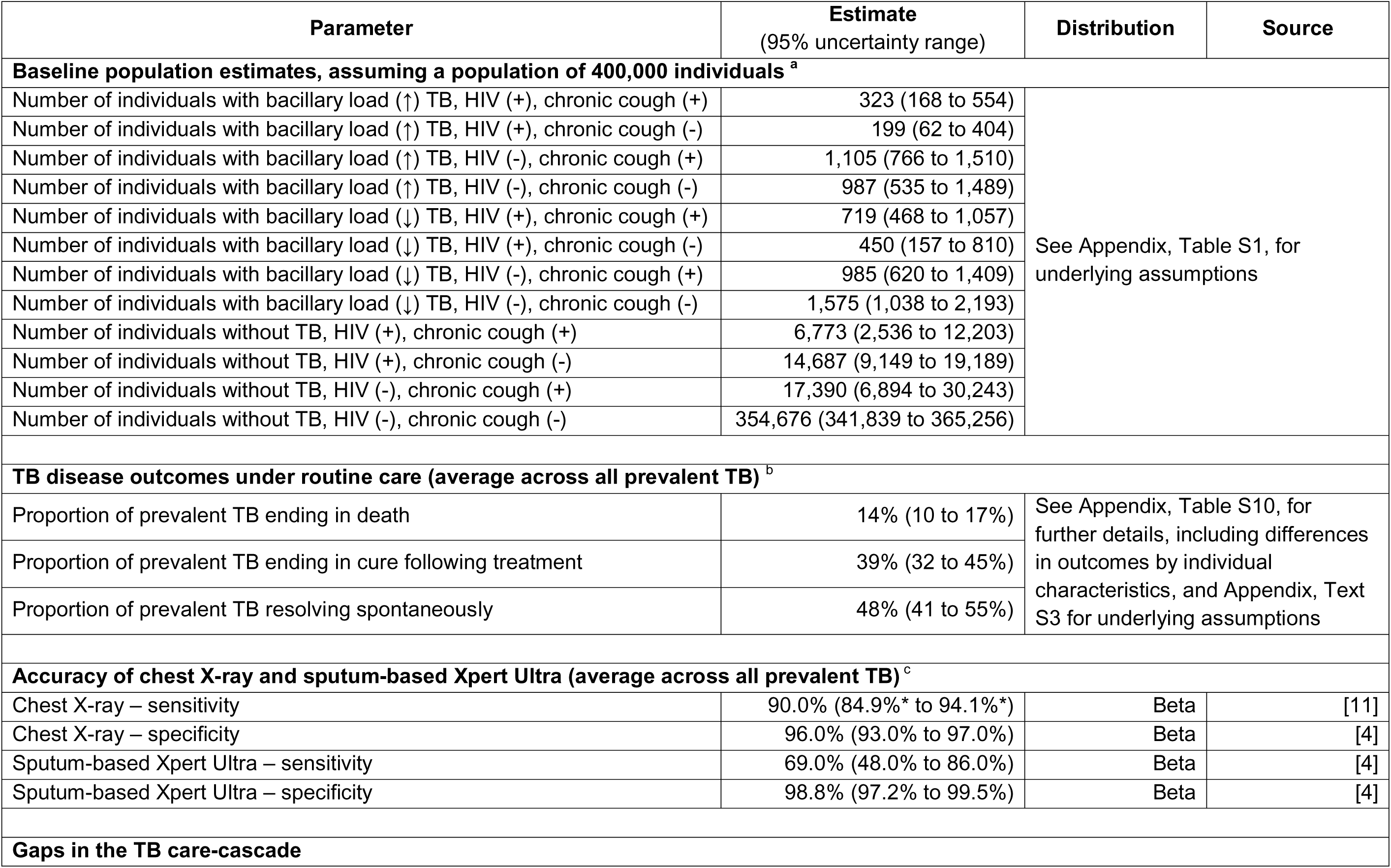
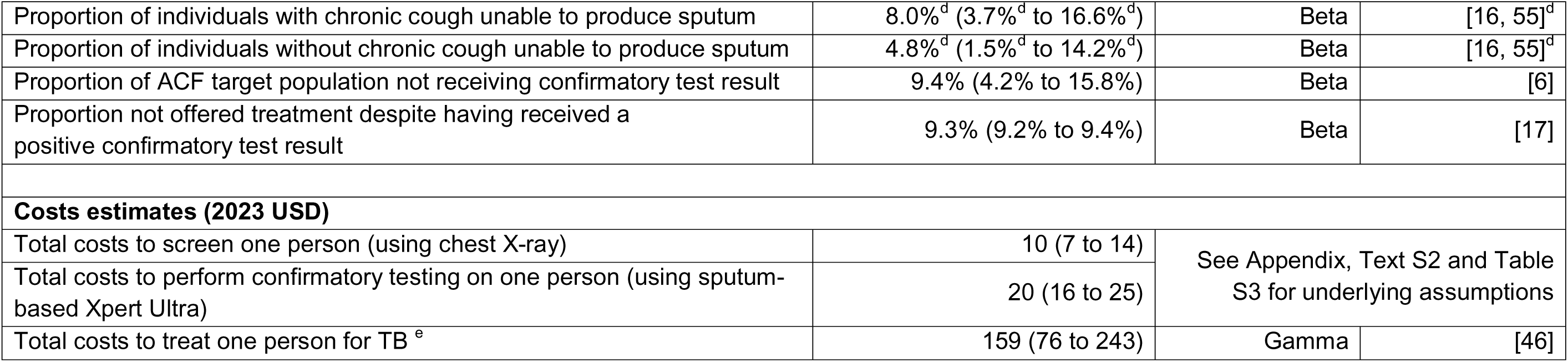
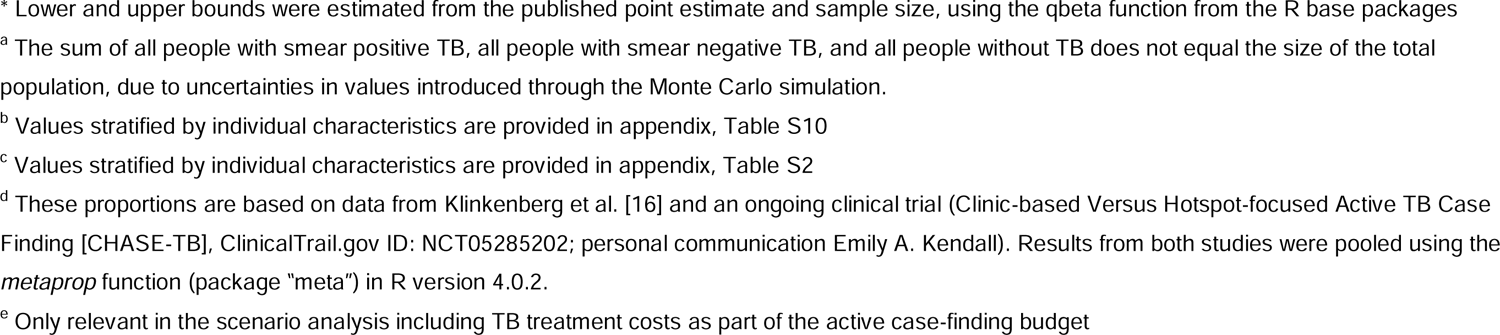
Key model parameters.

### Active case-finding

We model an ACF intervention consisting of community-based screening for TB, followed by confirmatory testing for those who screen positive. To reflect the current best available tools, we assume that the ACF intervention uses mobile chest X-ray to screen adults for TB, with immediate artificial intelligence-based reporting of results [4, 11, 15]. For people screening positive, we assume confirmatory testing to be immediately attempted, using sputum-based Xpert Ultra as the diagnostic test [5–7]. Individuals unable to produce sputum are excluded from confirmatory testing; sputum production is estimated based on systematic TB screening efforts modeling people with chronic cough as more likely to produce sputum than those without chronic cough [16]. We assume that referral to treatment services is attempted for all participants with positive confirmatory test results, but that some losses are incurred in notifying participants (for confirmatory tests not completed at the point of care) and in linkage to treatment. For participants who initiate treatment as a result of ACF, outcomes following treatment are allocated such that they are not worse (i.e., increased TB mortality or longer disease duration) than under routine care (Table 2 and Appendix, Text S1) [17–21].

### Costs of active case-finding and budget constraint

In order to compare various confirmatory test improvements, we consider an arbitrary limited budget of one million USD for the ACF effort, assuming that some of the target population will not be screened due to budget constraints. This budget includes screening and confirmatory testing costs, but we assume treatment costs for people diagnosed with TB to be budgeted separately; the latter assumption is modified in sensitivity analysis. For each modeled confirmatory test, we estimate the number of people who could be screened under the available screening and testing budget, based on the per-participant cost of the screening step (including staff, transport, and screening test costs), the proportion of participants who require confirmatory testing, and the per-person cost of confirmatory testing (Table 2 and Appendix, Text S2 and Table S3) [8, 22–28]. All costs are presented in 2023 USD (inflated using World Bank consumer price indices where applicable [29]) and from the healthcare system perspective.

### Confirmatory test improvements

Our primary comparison is between ACF utilizing Xpert Ultra on an expectorated sputum specimen (“baseline confirmatory testing”) [4, 9] and a series of ACF interventions with hypothetical improved confirmatory tests, each of which improves on the background of Xpert Ultra in one aspect (further details in Figure 1 and Table 1): (A) reduced test cost, (B) non-sputum specimen, (C) immediate turnaround, and (D) increased test sensitivity.

### Oral-swab based testing

One diagnostic testing strategy currently in development is oral-swab based testing. Oral swabs might be a more readily available specimen but result in reduced sensitivity (existing prototypes been have estimated to have sensitivities between 52% and 97% relative to sputum molecular testing [30, 31]). To reflect the potential impact of this testing method, we also project the minimal sensitivity required for oral-swab-based confirmatory testing to achieve at least the same epidemiological impact as the baseline confirmatory test. We assume that incremental false negatives occur in individuals with low bacillary load first [31] and that oral-swab sensitivity is independent of ability to expectorate except as mediated by the presence of chronic cough. To account for uncertainty in the proportion of people able to expectorate sputum for confirmatory testing, we perform these analyses at values of 93% (as in the primary analysis), 83%, and 73% for the proportion of people able to submit sputum (Appendix, Text S5).

### TB diagnosis and treatment under background routine care

For people with TB not included in ACF efforts – due to either budget constraints or losses along the ACF care cascade – outcomes of routine care are determined by our model.

Under routine care, an episode of TB can end before any treatment, either through death or through spontaneous resolution, or treatment can be initiated – in which case the disease course can end in either eventual cure or (for some whose treatment is unsuccessful) TB death after treatment (Figure 1).

For HIV-negative people, we estimate the competing probabilities of receiving treatment under routine care, or of dying or experiencing spontaneous TB resolution before any treatment, based on results from a published smear- and symptom-stratified model of TB natural history [32]. We use the same model’s estimates of disease duration to estimate the time that each person spends with untreated TB, separately estimating the cumulative amounts of time with high and with low bacillary loads (Appendix, Text S3 and Table S5). For HIV-positive individuals, we adjust these estimates using results from another model of TB disease courses that incorporated HIV-stratified estimates of TB notification and mortality; in the absence of data for Uganda, we apply estimates for Kenya [33–35] (Appendix, Text S3 and Table S5). Of note, the models underlying our work estimate a high proportion of smear-negative TB to spontaneously resolve prior to receiving treatment. Given that approximately 60% of people with prevalent TB in our modeled setting have a low bacillary load, our model also projects 48% (95% uncertainty range [UR]: 40-55%) of individuals with prevalent TB to spontaneously resolve prior to receiving treatment.

For people with TB who start treatment through routine care, the probability of an eventual outcome of cure versus TB death is projected based on a combination of treatment outcomes as reported to the World Health Organization plus clinical trial results to estimate relapse risk (Appendix, Text S3, Table S4, and Table S6). Estimates are of the eventual outcome of a TB episode, after any retreatments that may be required due to failure or relapse [18–21, 36, 37]. In estimating how relapse and failure contribute to the cumulative duration of TB, we assume that the average time spent with TB after an initial recurrence is equal to the average duration of TB prior to any treatment, after stratification by HIV status.

### Outcome measures and reporting

Our primary comparisons are of the health benefit of ACF, comparing ACF with the baseline confirmatory test to ACF that uses the improved confirmatory tests described above, when both are evaluated under the same constrained budget (Figure 1). We estimate the health benefit of ACF using the following measures:

1. the total number of people linked to treatment through ACF,
2. the number of TB-related deaths averted through ACF, and
3. the TB transmission potential averted through ACF (estimated as a difference in total high bacillary-load-equivalent person-months, adjusting for an estimated fourfold lower infectivity during time spent with low bacillary load [10]).

We then estimate the incremental change in each of these measures when the confirmatory test is improved, on absolute scales and relative to the impact achieved with the baseline test.

### Analysis and reporting

To capture uncertainty, we first simulate 10,000 iterations of the targeted community-based cohort of 400,000 adults and their disease courses under routine care; within each cohort, we then simulate all modeled diagnostic tests and perform pairwise comparisons. All parameters are independently sampled from beta (if bounded above by one) or gamma (if no upper bound) distributions reflecting the uncertainty in available primary data or published estimates (Table 2 and Appendix, Tables S1-6, with further details in Text S4). For each outcome, we report the median value across these simulated cohorts as the point estimate, with a 95% uncertainty range (UR) based on the 2.5^th^ and 97.5^th^ percentiles across all simulations. All analyses use R version 4.0.2 (R Foundation for Statistical Computing, Vienna, Austria). Ethical approval was not sought for this study as there was no human subject participation.

### Sensitivity and scenario analyses

We analyze one-way sensitivity of model results to setting-dependent variation in TB and HIV prevalence and diagnostic testing costs [15, 38–45] (Appendix Text S5 and Table S7). We also evaluate how results change in scenario analyses that (1) include treatment costs as part of the ACF budget (to assess the potential impact of improved confirmatory test specificity) [46], (2) assume that prevalent TB does not spontaneously resolve (while assuming that treatment initiations and deaths prior to treatment occur in the same ratio as in the base model), or (3) model costs [47] and accuracy of screening by chronic cough rather than by chest X-ray (Appendix Text S5 and Table S8).

## RESULTS

### Population estimates

Our model projected 6,421 (95% uncertainty range: 5,316-7,531) people with TB in the target population of 400,000 adults, reflecting the assumed 1.6% (1.3-1.9%) prevalence of TB. Of the people with TB in the target population, 2,716 (2,089-3,420; 42% [35-50%]) people were projected to receive treatment through routine care in absence of any ACF, and 2,466 (1,897-3,105; 91% [90-92%]) of those were projected to be cured. An estimated 873 (612-1,182; 14% [10-17%]) individuals were projected to die from TB in absence of ACF: 625 (377-915) before receiving treatment and 249 (183-328) afterwards. The cumulative duration of culture-positive and potentially infectious TB among this population in absence of ACF was 42,604 (31,409-56,519) infectivity adjusted person-months (Figure 2).

**Figure 2.**
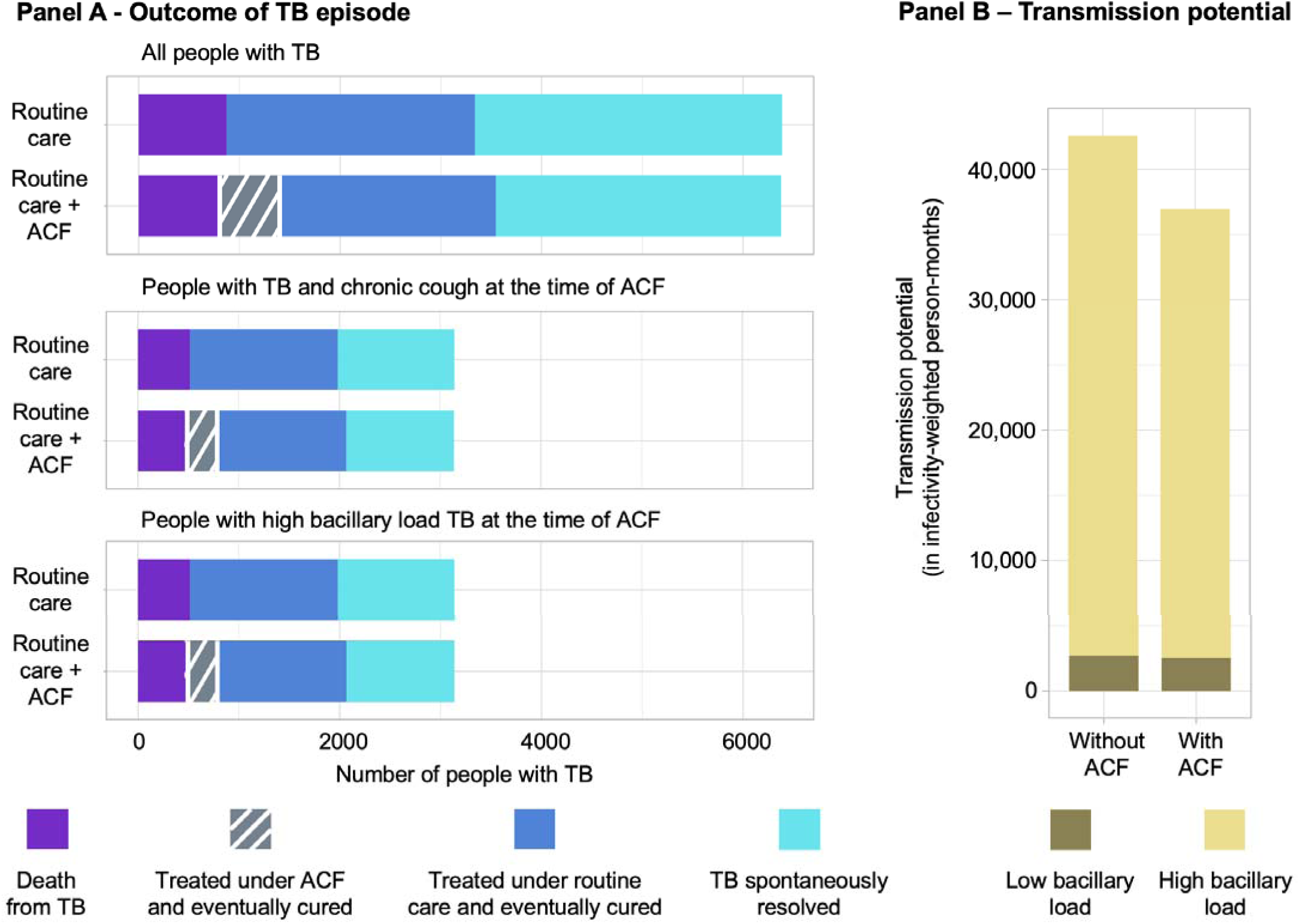
Projected epidemiological impact of a one-time active case-finding effort for tuberculosis in a high-burden population of 400,000 adults Bars depict the estimated epidemiological impact of active case-finding (ACF) for tuberculosis (TB) among adults using Xpert Ultra as confirmatory test, compared to a situation where no community-based screening is in place (assuming a setting similar to Uganda). Panel A shows the number of adults with TB (top), with TB and chronic cough (middle), and with high TB bacillary load (bottom) that are projected to die from TB (purple), be treated and cured from TB (grey or dark blue) and in whom TB might spontaneously resolve or be cured in subsequent treatment attempts (light blue). The dark blue refers to the number of people that are linked to treatment through routine care efforts, i.e., symptom-based passive case-finding, and grey to those receiving treatment under ACF efforts. Panel B shows the future transmission potential (in infectivity-weighted person months) resulting from people with prevalent TB with high bacillary load (i.e., where smear microscopy would be positive; light yellow) and low bacillary load (i.e., where smear microscopy would be negative; dark yellow) TB, when ACF is not in place (left) versus when community-based ACF is conducted (right).

### Community-based screening using Xpert Ultra

Accounting for a constrained budget of $1 million, and estimating costs of $10 ($7-14) per person for chest X-ray screening and $20 ($16-25) for confirmatory testing (Table 2 and Appendix Text S2), we estimated that the ACF budget would allow 83,808 (59,388-118,601; 21% [15-30%] of the target population) to undergo one-time TB screening. During this ACF intervention, TB would be detected in 718 (474-1,085) people, with 651 (429-983) of those started on treatment and 611 (403-924) eventually cured.

Of the people treated and cured as a result of ACF efforts with the baseline confirmatory test, 76 (39-132) were individuals who would have died had they not been detected through ACF. Thus, ACF reduced projected TB deaths in the target population to 796 (565-1,069), an 9% (6-13%) reduction compared to no ACF intervention. Furthermore, community-based ACF was projected to prevent 13% (9-18%) of future TB transmission potential from the people with prevalent TB in the target population: a reduction of 5,512 (3,402-8,818) infectivity-adjusted person-months (Figure 2).

### Impact of confirmatory test improvements

Of the test improvements modeled, increased sensitivity led to the largest incremental increase in TB treatment initiations (14% [4-26%] more people than through ACF with baseline confirmatory testing, 93 (29-180) additional people among the 83,808 individuals screened]). Other confirmatory test improvements increased the number of people linked to treatment through ACF by 11% (5-18%; immediate turn-around), 8% (5-12%; non-sputum specimen), and 6% (2-22%; reduced test costs).

By contrast, when considering mortality outcomes, the largest impact was found with immediate turn-around (11% [5-18%] more deaths averted than through ACF with baseline confirmatory testing; 8 [3–17] incremental deaths averted), followed by using a non-sputum specimen (8% [4-12%] increase), reduced test costs (6% [2-22%] increase), and increased test sensitivity (5% [2-11%] increase). Relative results for averted transmission potential were similar to those for deaths averted (Figure 3 and Table 3).

**Figure 3.**
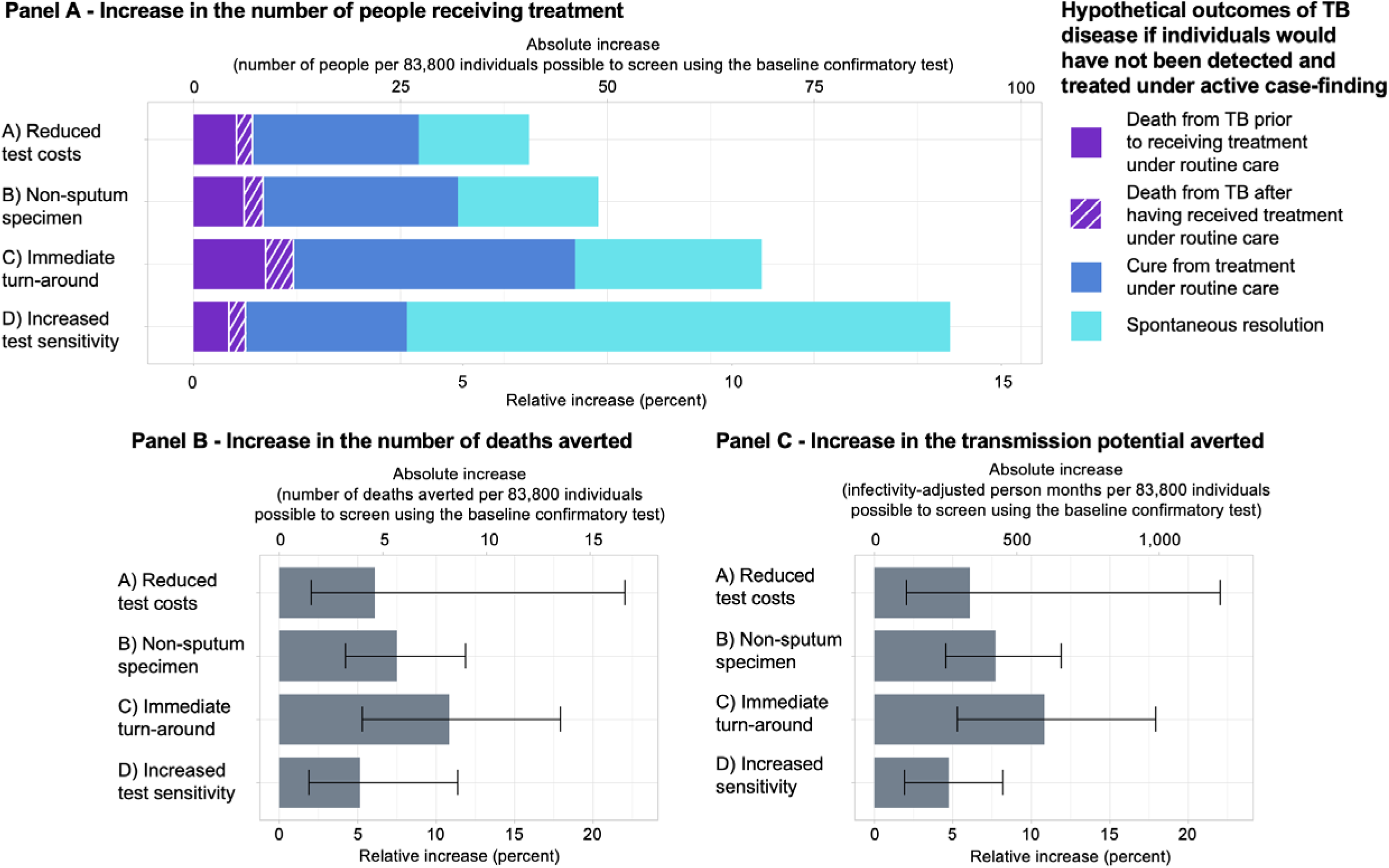
Epidemiological effect of an improved test to confirm tuberculosis, when used in an active case-finding campaign with a budget of 1 million US dollars Bars show the potential epidemiological benefit of ACF using hypothetical improved confirmatory tests when screening a target population of 400,000 people for tuberculosis (TB) compared to conducting the same ACF efforts using Xpert Ultra to confirm a positive screening test result. An estimated 82,400 individuals could be screened under the allocated budget in the baseline comparator scenario. Test improvements considered are: (1) Increase in test sensitivity (from 69.0% to 80.0%; purple bar), using a non-sputum respiratory specimen type (increasing confirmatory specimen production from 93% to 100% for those eligible), (3) point-of-care testing (increasing the proportion of receiving the confirmatory test result from 91% to 100%), and (4) reduced costs (from $20 to $10 per confirmatory test). Panel A depicts the increase in the number of people with TB diagnosed and treated under ACF efforts when each of the named confirmatory test improvements is used compared to ACF utilizing the baseline confirmatory test. Herein, the purple area of the bars refers to people with TB who would have died in the absence of community-based screening, the dark blue area to people with TB who would have received treatment and been eventually cured even in the absence of ACF, and the light blue area to people in whom TB would have spontaneously resolved prior to receiving any treatment. Panel B shows the estimated reduction in TB mortality (number of TB-related deaths) and Panel C the projected reduction in TB transmission (infectivity-adjusted person-months) resulting from each of the test improvements.

**Table 3.**
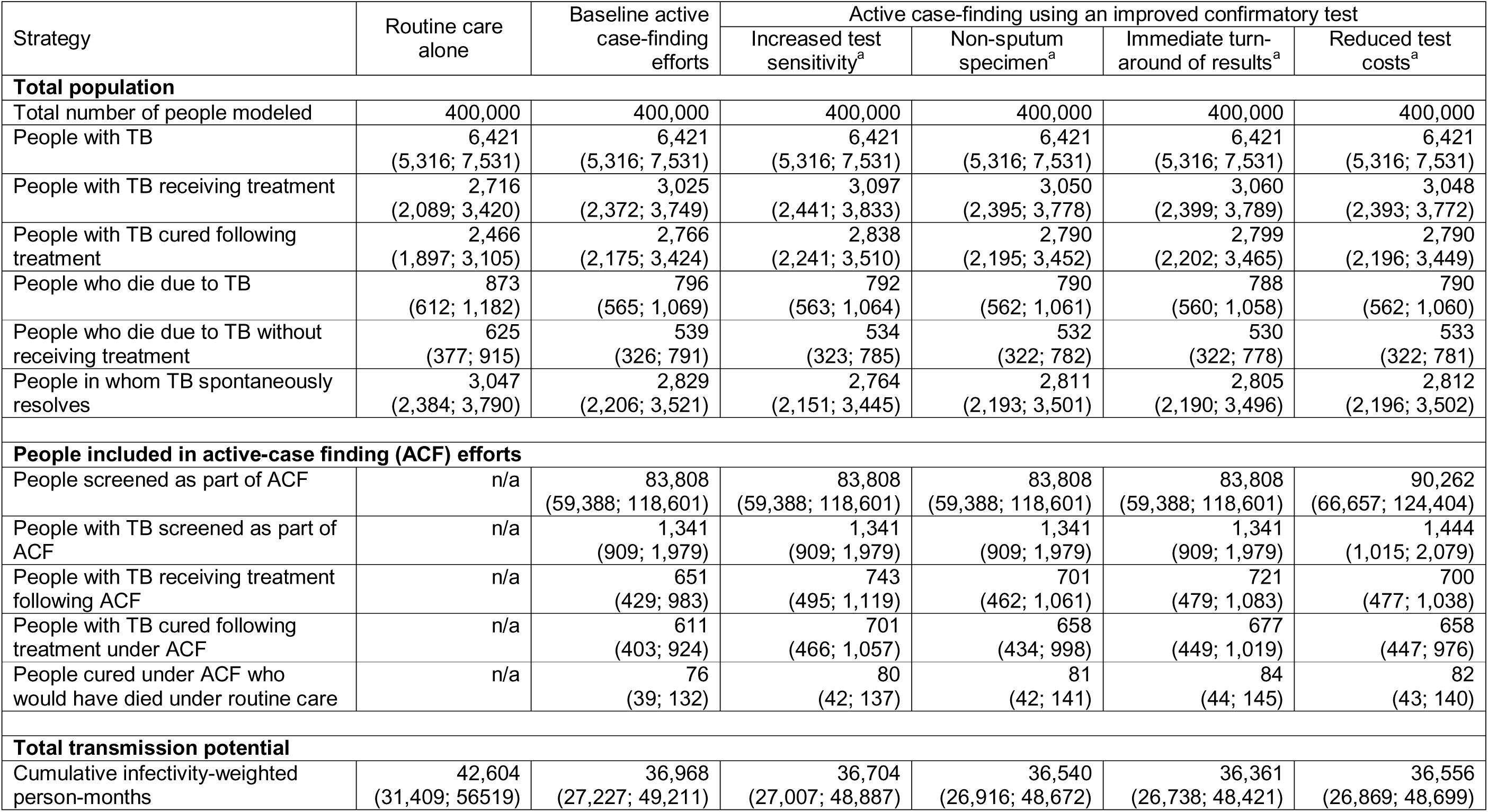
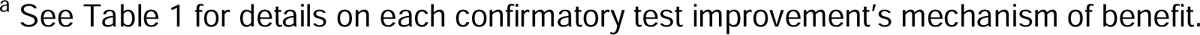
Epidemiological outcomes under active case finding for tuberculosis, using alternative tests to confirm positive screening results.

### Oral-swab based testing

For an oral-swab based test with reduced sensitivity to match the mortality impact of baseline sputum confirmatory testing (assuming 93% of the study population to be able to produce sputum), we estimated that oral-swab based testing would need to be at least 78% [77-82%] as sensitive as the sputum-based confirmatory test. This sensitivity requirement fell to 63% [62-67%] and 52% [51-56%] when assuming lower (83% and 73%, respectively) sputum production. Results were similar when considering transmission potential averted as a metric (Figure S1).

### Sensitivity and scenario analysis

Neither variation of TB or HIV burden or tests costs in one-way sensitivity analyses (Text S6 and Figure S2) nor consideration of treatment costs as part of the ACF budget (Text S6 and Figure S4) materially affected the relative importance of different confirmatory test improvements. When including treatment costs, specificity also remained less influential than other confirmatory test characteristics (Figure S4). However, when spontaneous TB resolution was eliminated from the model, increased sensitivity became the confirmatory test characteristic leading to the greatest reductions in TB mortality and transmission potential (Text S6 and Figure S3). Assuming chronic-cough-based rather than radiographic screening (i.e., screening at lower cost, sensitivity and specificity), the reach of screening increased to 279,271 individuals (129,894-414,818; baseline scenario: 83,808 [59,388–118,601]), thereby tripling the number of deaths averted to 238 (95-468; baseline scenario: 76 [39–132]) and infectivity-adjusted person-months averted to 17,871 (7,949-31,033; baseline scenario: 5,512 [3,402–8,818]). When screening by chronic cough, test costs became the most relevant confirmatory test improvement, with reduced confirmatory test costs leading to a 23% (9-56%) increase in incremental TB deaths and incremental TB transmission potential averted (Figure S5).

## DISCUSSION

This modeling analysis evaluates the potential epidemiological impact of improvements to current tools for confirmatory testing in community-based screening (i.e., ACF) for TB. Although accuracy is often prioritized in development of TB diagnostic tests, we predict the use of a highly-available specimen type and immediacy of turn-around to have greater impact on mortality and transmission potential, when considering a confirmatory testing use case during TB screening of high-TB-prevalence community. However, none of these improvements in isolation increased the mortality or transmission impact of ACF by more than 11%; thus, a greater variety of enhancements to feasibility and effectiveness are needed if ACF is to play a major role in meeting WHO End TB targets [48].

Our results indicate that an increase in confirmatory test sensitivity (from 69% to 80%) would result in the largest increase in the number of people diagnosed and treated through ACF, but would yield only small benefit in terms of reducing TB mortality and transmission. This discrepancy occurs because the people incrementally diagnosed by a more sensitive test would have lower bacillary loads; besides having lower current infectivity [10, 49], survival data from the pre-antibiotic data suggest such individuals have lower TB mortality risk [50], and models also translate this into less cumulative future transmission [32]. By contrast, reducing operational barriers to confirmatory testing, e.g., with a point-of-care test or a non-sputum sample type, would enable detection of additional people with high bacillary load (i.e., more fatal and more transmissible) TB. Hence, focusing efforts on reducing operational barriers in confirmatory testing might achieve greater population health benefits than further increasing test sensitivity. This is consistent with recent modeling showing that greater accessibility may be more impactful than high sensitivity in clinical diagnostic settings [51] – strengthening the case for focusing diagnostics development efforts on improving cost and ease of use.

The relative importance of sensitivity or cost characteristics could change under certain circumstances. Our primary analyses assume that community-based active case-finding would use some of the most accurate screening tools currently available, i.e., chest X-ray. However, if a less specific screening tool, such as symptom screening [11], is used, more people will require confirmatory testing, and confirmatory testing will comprise a greater proportion of the overall ACF budget. As seen in our scenario analysis, this increases the importance of confirmatory test costs relative to operational improvements in determining intervention reach and impact. In addition, certain test improvements, such as non-sputum-based testing, might require counterbalancing reductions in sensitivity. For example, we estimate that oral-swab-based testing – the currently most promising technique for complementing sputum testing – would require at least 63-78% the sensitivity of sputum-based testing to achieve a similar population health benefit in a setting where 83-93% of individuals are able to produce sputum. Most recent studies of tongue swab testing among symptomatic individuals presenting for care suggest sensitivities at least this high [31, 52]. If sensitivity at this level can be achieved in the context of community-based screening, oral-swab samples might provide a valuable addition to sputum-based testing for confirmatory testing in ACF interventions.

We estimated that isolated improvements to confirmatory tests were likely to increase the overall mortality and transmission impact of ACF by no more than 11%. In the context of population-wide systematic screening, this incremental benefit could be comparable to, for example, the estimated benefit of hypothetically improved TB treatment regimens (assuming 99% efficacy and 2 months treatment duration) [19]. Given expected synergies between different test improvements (e.g., if a non-sputum specimen were available, more individuals could complete confirmatory testing, further increasing the benefit of immediate results turn-around), the combined effect of improving multiple test characteristics could exceed the sum of their individual effects. Nonetheless, to optimally enhance the impact and cost-effectiveness of community-based active case-finding, other improvements than confirmatory testing should be pursued as well; possibilities include better tools for identifying high-risk populations, simpler and more affordable screening tests, or efforts to combine ACF for TB with other health screening activities [53]. Thus, while changes through improved confirmatory testing would not be transformative, they could meaningfully increase the effectiveness and cost-effectiveness of ACF, especially if multiple improvements could be combined.

Our results are limited by simplifying assumptions in our model’s representation of TB disease states, case-finding interventions, and treatment outcomes. Because it is not ethical to perform observational studies of the untreated TB disease course, disease durations and outcomes must be inferred from cross-sectional and historical data. Thus, our estimates of the outcomes of prevalent TB under routine care, which we based on prior modeling analyses, are subject to those models’ uncertainties, including data limitations (e.g., on HIV-associated TB), reliance on historical microbiological classifications, and uncertain accuracy of country-level TB notification and mortality tabulations [32, 33]. Moreover, we have focused on how improvements to diagnostic tests would affect the outcomes of community-wide screening, but it is likely that those improvements would also enhance other TB interventions (e.g., contact screening and prevention) in ways we have not modelled. Furthermore, we dichotomize more nuanced population characteristics such as HIV severity, presence of chronic cough, and bacillary load. Lastly, our modeling of treatment outcomes is simplified by not explicitly modeling re-treatments, drug resistance, or relationships between the timing of diagnosis and treatment outcomes.

In conclusion, we found that reducing operational barriers for confirmatory TB testing – such as changing to a non-sputum specimen or facilitating immediate turn-around of test results – is likely to lead to greater impact on TB transmission and mortality in the context of community-based ACF than increasing sensitivity. Since improvements in confirmatory tests are likely to have modest epidemiological impact in isolation, other measures to improve ACF should be explored as well.

## ETHICS APPROVAL AND CONSENT TO PARTICIPATE

Not applicable.

## CONSENT FOR PUBLICATION

Not applicable.

## AVAILABILITY OF DATA AND MATERIALS

All data generated or analyzed during this study are included in this published article (and its supplementary information files).

## AUTHORS’ CONTRIBUTIONS

S. S., F. M. M., C. M. D., D. W. D., and E. A. K. conceived the study. L. E. B. and E.A.K. performed the analysis, wrote a first draft of the manuscript, and finalized the manuscript based on co-authors comments. T.S.R. provided simulation-model results as inputs to the TB outcomes decision-tree model, and advised on the modeling approach. W.W., D.J.C., G.T., and A.C. provided substantial comments to a first draft of the analyses. All authors reviewed the manuscript and analyses and approved the final version for submission.

## Data Availability

All data produced in the present work are contained in the manuscript

## ACKNOWELEDGEMENTS

We acknowledge support from the Else Kröner-Fresenius-Stiftung within the Heidelberg Graduate School of Global Health and from the German Academic Exchange Foundation to L. E. B.

## REFERENCES

1. World Health Organization. Global Tuberculosis Report 2023. Geneva (Switzerland); 2023.

2. Dowdy DW, Basu S, Andrews JR. Is passive diagnosis enough? The impact of subclinical disease on diagnostic strategies for tuberculosis. Am J Respir Crit Care Med. 2013;187(5):543–51.

3. Fenta MD, Ogundijo OA, Warsame AAA, Belay AG. Facilitators and barriers to tuberculosis active case findings in low- and middle-income countries: a systematic review of qualitative research. BMC Infect Dis. 2023;23(1):515.

4. World Health Organization. WHO consolidated guidelines on tuberculosis. Module 2: Screening. Geneva (Switzerland); 2021.

5. Floyd S, Klinkenberg E, de Haas P, Kosloff B, Gachie T, Dodd PJ, et al. Optimising Xpert-Ultra and culture testing to reliably measure tuberculosis prevalence in the community: findings from surveys in Zambia and South Africa. BMJ Open. 2022;12(6):e058195.

6. Kendall EA, Kitonsa PJ, Nalutaaya A, Erisa KC, Mukiibi J, Nakasolya O, et al. The Spectrum of Tuberculosis Disease in an Urban Ugandan Community and Its Health Facilities. Clin Infect Dis. 2021;72(12):e1035–e43.

7. Puma D, Yuen CM, Millones AK, Brooks MB, Jimenez J, Calderon RI, et al. Sensitivity of Various Case Detection Algorithms for Community-based Tuberculosis Screening. Clin Infect Dis. 2023;76(3):e987–e9.

8. Hsiang E, Little KM, Haguma P, Hanrahan CF, Katamba A, Cattamanchi A, et al. Higher cost of implementing Xpert((R)) MTB/RIF in Ugandan peripheral settings: implications for cost-effectiveness. Int J Tuberc Lung Dis. 2016;20(9):1212–8.

9. Dorman SE, Schumacher SG, Alland D, Nabeta P, Armstrong DT, King B, et al. Xpert MTB/RIF Ultra for detection of Mycobacterium tuberculosis and rifampicin resistance: a prospective multicentre diagnostic accuracy study. Lancet Infect Dis. 2018;18(1):76–84.

10. Tostmann A, Kik SV, Kalisvaart NA, Sebek MM, Verver S, Boeree MJ, et al. Tuberculosis transmission by patients with smear-negative pulmonary tuberculosis in a large cohort in the Netherlands. Clin Infect Dis. 2008;47(9):1135–42.

11. The Republic of Uganda. The Uganda National Tuberculosis Prevalence Survey, 2014-2015 Survey Report. 2017.

12. The Republic of Uganda. Uganda population-based HIV impact assessment. 2022.

13. Sekandi JN, List J, Luzze H, Yin XP, Dobbin K, Corso PS, et al. Yield of undetected tuberculosis and human immunodeficiency virus coinfection from active case finding in urban Uganda. Int J Tuberc Lung Dis. 2014;18(1):13–9.

14. World Health Organization. Global Tuberculosis Report 2014. Geneva; 2014.

15. Republic of South Africa National Department of Health. The First National TB Prevalence Survey - South Africa 2018. Pretoria (South Africa); 2021.

16. Klinkenberg E, Floyd S, Shanaube K, Mureithi L, Gachie T, de Haas P, et al. Tuberculosis prevalence after 4 years of population-wide systematic TB symptom screening and universal testing and treatment for HIV in the HPTN 071 (PopART) community-randomised trial in Zambia and South Africa: A cross-sectional survey (TREATS). PLoS Mwed. 2023;20(9):e1004278.

17. Lungu P, Kerkhoff AD, Kasapo CC, Mzyece J, Nyimbili S, Chimzizi R, et al. Tuberculosis care cascade in Zambia - identifying the gaps in order to improve outcomes: a population-based analysis. BMJ Open. 2021;11(8):e044867.

18. World Health Organization. Tuberculosis data, CVS files to download Geneva (Switzerland)2023 [Available from: https://www.who.int/teams/global-tuberculosis-programme/data.

19. Kendall EA, Shrestha S, Cohen T, Nuermberger E, Dooley KE, Gonzalez-Angulo L, et al. Priority-Setting for Novel Drug Regimens to Treat Tuberculosis: An Epidemiologic Model. PLoS Med. 2017;14(1):e1002202.

20. Knight GM, Gomez GB, Dodd PJ, Dowdy D, Zwerling A, Wells WA, et al. The Impact and Cost-Effectiveness of a Four-Month Regimen for First-Line Treatment of Active Tuberculosis in South Africa. PLoS One. 2015;10(12):e0145796.

21. Velayutham B, Chadha VK, Singla N, Narang P, Gangadhar Rao V, Nair S, et al. Recurrence of tuberculosis among newly diagnosed sputum positive pulmonary tuberculosis patients treated under the Revised National Tuberculosis Control Programme, India: A multi-centric prospective study. PLoS One. 2018;13(7):e0200150.

22. Baik Y, Nakasolya O, Isooba D, Mukiibi J, Kitonsa PJ, Erisa KC, et al. Cost to perform door-to-door universal sputum screening for TB in a high-burden community. Int J Tuberc Lung Dis. 2023;27(3):195–201.

23. Stop TB Partnership. Artificial intelligence-powered computer-aided (CAD) software 2022 [Available from: https://www.stoptb.org/introducing-new-tools-project/artificial-intelligence-powered-computer-aided-detection-cad-software.

24. Stop TB Partnership. Diagnostics, medical devices & other health products catalog 2023 [Available from: https://www.stoptb.org/sites/default/files/gdf_diagnostics_medical_devices_other_health_products_catalog_0.pdf.

25. Jiji Vehicels. Toyota Buses & Microbuses in Uganda 2023 [Available from: https://jiji.ug/buses/toyota.

26. Global Petrol Prices. Uganda Gasoline prices, litre, 21-Aug-2023 2023 [Available from: https://www.globalpetrolprices.com/Uganda/gasoline_prices/#:~:text=Uganda%3A%20The%20price%20of%20octane,see%20the%20prices%20in%20gallons.

27. The Global Fund. Global Fund, Stop TB Partnership and USAID Announce New Collaboration with Danaher to Reduce Price and Increase Access to Cepheid’s TB Test. 2023.

28. Thompson RR, Nalugwa T, Oyuku D, Tucker A, Nantale M, Nakaweesa A, et al. Multicomponent strategy with decentralised molecular testing for tuberculosis in Uganda: a cost and cost-effectiveness analysis. Lancet Glob Health. 2023;11(2):e278–e86.

29. The World Bank. Inflation, consumer prices (annual %) - United States 2023 [Available from: https://data.worldbank.org/indicator/FP.CPI.TOTL.ZG?locations=US.

30. Church EC, Steingart KR, Cangelosi GA, Ruhwald M, Kohli M, Shapiro AE. Oral swabs with a rapid molecular diagnostic test for pulmonary tuberculosis in adults and children: a systematic review. Lancet Glob Health. 2024;12(1):e45–e54.

31. Steadman A, Andama A, Ball A, Mukwatamundu J, Khimani K, Mochizuki T, et al. New manual qPCR assay validated on tongue swabs collected and processed in Uganda shows sensitivity that rivals sputum-based molecular TB diagnostics. Clin Infect Dis. 2024.

32. Ryckman TS, Dowdy DW, Kendall EA. Infectious and clinical tuberculosis trajectories: Bayesian modeling with case finding implications. Proc Natl Acad Sci U S A. 2022;119(52):e2211045119.

33. Ku CC, MacPherson P, Khundi M, Nzawa Soko RH, Feasey HRA, Nliwasa M, et al. Durations of asymptomatic, symptomatic, and care-seeking phases of tuberculosis disease with a Bayesian analysis of prevalence survey and notification data. BMC Med. 2021;19(1):298.

34. UNAIDS. AIDSinfo – Global data on HIV epidemic and response Geneva (Switzerland)2023 [Available from: https://aidsinfo.unaids.org.

35. United Nations. Crude death rate per country. Manhattan (New York, United States); 2023.

36. Akessa GM, Tadesse M, Abeb G. Survival Analysis of Loss to Follow-Up Treatment among Tuberculosis Patients at Jimma University Specialized Hospital, Jimma, Southwest Ethiopia. International Journal of Statistical Mechanics. 2015;2015.

37. Fox GJ, Nguyen VN, Dinh NS, Nghiem LPH, Le TNA, Nguyen TA, et al. Post-treatment Mortality Among Patients With Tuberculosis: A Prospective Cohort Study of 10 964 Patients in Vietnam. Clin Infect Dis. 2019;68(8):1359–66.

38. AIDS Healthcare Foundation. The fifth South African national HIV prevalence, incidence, behaviour and communication survey. 2018.

39. Ministry of Health & Family Welfare - Government of India. India Tuberculosis Report. 2022.

40. Muniyandi M, Lavanya J, Karikalan N, Saravanan B, Senthil S, Selvaraju S, et al. Estimating TB diagnostic costs incurred under the National Tuberculosis Elimination Programme: a costing study from Tamil Nadu, South India. Int Health. 2021;13(6):536–44.

41. National AIDS Control Organization & ICMR-National Institute of Medical Statistics. India HIV Estimates 2021: Fact Sheet. New Delhi, India: Ministry of Health and Family Welfare, Government of India; 2022.

42. Pooran A, Theron G, Zijenah L, Chanda D, Clowes P, Mwenge L, et al. Point of care Xpert MTB/RIF versus smear microscopy for tuberculosis diagnosis in southern African primary care clinics: a multicentre economic evaluation. Lancet Glob Health. 2019;7(6):e798–e807.

43. Sathiyamoorthy R, Kalaivani M, Aggarwal P, Gupta SK. Prevalence of pulmonary tuberculosis in India: A systematic review and meta-analysis. Lung India. 2020;37(1):45–52.

44. Cambodia Ministry of Health. Report of the second national tuberculosis prevalence survey, 2011. Phnom Penh; 2012.

45. Federal Republic of Nigeria. First National TB Prevalence Survey 2012, Nigeria. Abuja, Nigeria; 2012.

46. Kairu A, Orangi S, Oyando R, Kabia E, Nguhiu P, Ong Ang OJ, et al. Cost of TB services in healthcare facilities in Kenya (No 3). Int J Tuberc Lung Dis. 2021;25(12):1028–34.

47. Machekera SM, Wilkinson E, Hinderaker SG, Mabhala M, Zishiri C, Ncube RT, et al. A comparison of the yield and relative cost of active tuberculosis case-finding algorithms in Zimbabwe. Public Health Action. 2019;9(2):63–8.

48. World Health Organization. The End TB strategy. Geneva; 2015.

49. Garcia LS, Costa AG, Araujo-Pereira M, Spener-Gomes R, Aguiar AF, Souza AB, et al. The Xpert(R) MTB/RIF cycle threshold value predicts M. tuberculosis transmission to close contacts in a Brazilian prospective multicenter cohort. Clin Infect Dis. 2024.

50. Tiemersma EW, van der Werf MJ, Borgdorff MW, Williams BG, Nagelkerke NJ. Natural history of tuberculosis: duration and fatality of untreated pulmonary tuberculosis in HIV negative patients: a systematic review. PLoS One. 2011;6(4):e17601.

51. Nooy A, Ockhuisen T, Korobitsyn A, Khan SA, Ruhwald M, Ismail N, et al. Trade-offs between clinical performance and test accessibility in tuberculosis diagnosis: a multi-country modelling approach for target product profile development. Lancet Glob Health. 2024;12(7):e1139–e48.

52. Andama A, Whitman GR, Crowder R, Reza TF, Jaganath D, Mulondo J, et al. Accuracy of Tongue Swab Testing Using Xpert MTB-RIF Ultra for Tuberculosis Diagnosis. J Clin Microbiol. 2022;60(7):e0042122.

53. Hill PC, Dye C, Viney K, Tabutoa K, Kienene T, Bissell K, et al. Mass treatment to eliminate tuberculosis from an island population. Int J Tuberc Lung Dis. 2014;18(8):899–904.

54. World Health Organization. High-priority target product profiles for new tuberculosis diagnostics: report of a consensus meeting. Geneva; 2014.

55. Moyo S, Ismail F, Van der Walt M, Ismail N, Mkhondo N, Dlamini S, et al. Prevalence of bacteriologically confirmed pulmonary tuberculosis in South Africa, 2017-19: a multistage, cluster-based, cross-sectional survey. Lancet Infect Dis. 2022;22(8):1172–80.

