## Supplemental File for "Importance of confirmatory test characteristics in optimizing community-based screening for tuberculosis: An epidemiological modeling analysis"

**APPENDIX**

[Table S4 – Estimated untreated TB disease durations under routine care for HIV-negative individuals, drawn from simulations by Ryckman et al [17]	27](#_Toc171892582)

### Text S1 – Tuberculosis diagnosis and treatment under active case-finding.

We project the impact of a one-time active case-finding (ACF) intervention on TB deaths and transmission in a Uganda-like, high-TB-prevalence setting, where passive, symptom-based case-finding is continuously available (“routine care”; Text S3). We assume the ACF intervention to last for two years, during which all participants are screened at most once with chest X-ray. A positive chest X-ray result is followed by immediately attempting confirmatory testing. Individuals who are also positive on the confirmatory test, and who can be contacted to deliver their results, are offered treatment for TB.

In estimating the deaths and transmission averted by ACF, we treat the target population as cross-sectional, even though the timing of the ACF intervention might not be simultaneous for all individuals. Because the local epidemic is assumed to be near steady state, and the timing of the ACF intervention for any given person is random relative to their disease course, the collective outcome of prevalent TB in the intervention’s target population is expected to be the same as that of a cross-sectional sample of the same size, who receive such an intervention simultaneously or are followed from a single point in time in absence of any such intervention.

Below, we first describe the process of screening and confirmatory testing as represented in our model, as well as any potential loss to follow up modeled as part of this diagnostic cascade. Afterwards, we outline how treatment outcomes were determined for any prevalent TB detected and linked to treatment through ACF.

#### 1.1 Screening and confirmatory testing

All individuals included in ACF receive a chest X-ray to screen for signs of TB. Chest X-ray sensitivity is assumed to be higher for people with high bacillary load (compared to low bacillary load), but for a given bacillary load it does not vary by HIV or the presence of chronic cough (i.e., cough for >2 weeks) [1, 2]. We assume chest X-ray specificity to be lower in people with HIV (because HIV can lead to non-TB-related abnormalities such as adenopathy or pulmonary infiltrates, potentially causing false-positive chest X-ray results) and in people with chronic cough (because other symptomatic respiratory conditions may cause a false-positive chest X-ray result) [1, 3] (main manuscript, Table 2).

For those screening positive on chest X-ray, confirmatory testing is immediately attempted. In the baseline scenario, Xpert Ultra MTB/RIF (Cepheid; Sunnyvale, CA, United States; “Xpert Ultra”) on an expectorated sputum specimen is used as the confirmatory test. We compare this Xpert Ultra standard of care to a hypothetical improved confirmatory test (as described in the manuscript’s methods section and Table 1). We assume that improved confirmatory tests will be bacteriologic tests on respiratory-tract specimens (e.g., the novel tests considered could include tongue swab molecular tests, but not a blood-based biomarker or urine test that correlates poorly with sputum bacillary burden), and thus for both Xpert Ultra and novel confirmatory tests, sensitivity is assumed to be higher for TB with high bacillary load (compared to low bacillary load) [2, 4]. Specificity of confirmatory tests is assumed to be invariant across the entire population [2] (main manuscript, Table 2).

We assume that all participants in the ACF efforts receive chest X-ray screening, but we consider potential loss to follow up at the confirmatory testing step due to inability to produce sufficient sputum. Ability to produce sputum is modeled as independent of HIV and bacillary load, but higher in individuals with chronic cough than in those without. We estimate respective proportions using data from an ongoing clinical trial (Clinic-based Versus Hotspot-focused Active TB Case Finding [CHASE-TB], ClinicalTrail.gov ID: NCT05285202) conducting chest X-ray screening with sputum Xpert Ultra confirmatory testing in peri-urban settings in Uganda (personal communication, Emily A. Kendall) as well as data from a survey of TB prevalence in urban and peri-urban communities in South Africa and Zambia [5]. Results from both studies were pooled using the *metaprop* function (from the package *meta*) in R version 4.2.2.

In addition, our model includes a potential for pretreatment LTFU due to not receiving the confirmatory test result (because of the need to contact patients with the results of tests that cannot be completed during the initial screening encounter) [6] (main manuscript, Table 2). An additional risk of pre-treatment LTFU, after receiving a positive Xpert Ultra result, is modeled for all participants who test positive [7] (main manuscript, Table 2).

#### 1.2 Treatment outcomes for prevalent TB detected and linked to treatment through ACF

In the context of the ACF intervention modeled here, we assume treatment is offered only to people with a positive Xpert test result. The possibility of empiric treatment during the ACF intervention is not included because it is a community-based intervention targeting people with no or mild symptoms. Thus, the non-clinician community health workers likely to implement such an intervention would not be trained to make judgment calls about treatment of TB in people who do not meet the criteria of standard diagnostic algorithms.

In modeling treatment outcomes for TB that was treated as a result of the ACF intervention, we assumed (for simplicity's sake, given absence of evidence to the contrary) that for people who would have been treated eventually under routine care (Text S3), their treatment outcomes were no different when diagnosed and treated earlier through ACF than they would have been if treated later through routine care. For people whose TB outcome under routine care would have been spontaneous resolution prior to treatment, we assumed treatment under ACF to always result in cure. We assumed people who would have experienced death prior to TB treatment under routine care, when detected and linked to treatment through ACF, to have the same treatment outcome probabilities as in Text S3.

Text S2 – Estimation of the cost for TB testing and treatment.

We compute the costs of active case-finding (ACF) to be the sum of per-test chest X-ray costs (i.e., screening) and per-test costs of Xpert Ultra (i.e., confirmatory testing costs). Due to large testing volumes, we consider all costs as variable costs. Assuming that all treatment costs come from other budgetary sources, we do not consider the costs for treating people diagnosed under community-based screening efforts to be part of the ACF costs (but perform a scenario analysis that includes treatment costs as part of the ACF budget).

In estimating chest X-ray costs, we account for equipment costs and staff costs. The largest proportion of equipment costs result from the X-ray machines (budgeted as Delft Light packages, plus costs for installation and two-year warranty) [8]. Considering the ACF intervention to last for two years, we assume 50% depreciation for the chest X-ray machines over this time frame. Further, costs for software to read chest X-ray results (CAD4TB; including costs for installation, a box to let the software run offline, and two-year support) and a solar panel to provide electricity to machine and software are included [9]. Also, we take into account costs for a mini-van to transport the chest X-ray machine. Specifically, we depict costs of $20,000 for buying a used Toyota HiAce (requiring ca. 7.5 liter fuel per 100km), with 30% depreciation over two years, and fuel costs of $1.30/liter, assuming the mini-van to drive 40,000km over the two years [10, 11].

In estimating staff costs, we assume a screening team consisting of one managerial-level staff member and one entry-level staff member, as well as one driver for the mini-van (also budgeted as entry-level staff) [12]. Estimating staff to work on average 50 weeks per year and five days per week, screening 50 people per day, we project each of these “screening teams” to be able to screen 25,000 people within two years.

In estimating confirmatory testing costs, we use values from the USAID-Cepheid buy-down agreement and in-country costing-studies to estimate the cartridge and non-cartridge costs of Xpert Ultra [13, 14]. For the sensitivity analysis where treatment costs are included as part of the ACF budget, we use average per-unit costs for first-line treatment in Kenya (estimated following a bottom-up approach), as no similar parameters were available for Uganda in recent studies [15]. All costs are presented in 2023 USD from the healthcare system perspective. When applicable, costs were inflated to 2023 USD using the United State’s World Bank gross domestic product (GDP) deflator [16]. All described costs are summarized in the main manuscript, Table 2**.**

### Text S3 – Tuberculosis diagnosis and treatment under routine care.

We assume our model to be placed in a setting where, in the absence of ACF efforts, passive case-finding occurs on a continuous basis (“routine care”). Possible outcomes of a TB episode are spontaneous resolution before attempted treatment, death due to TB either before or after attempted treatment, or cure following treatment (possibly requiring more than one course of treatment to achieve). The probability of each of these outcomes under a scenario of routine care only, as well as the corresponding duration of TB disease until one of those outcomes is reached, are estimated through three steps:

1. We use extrapolations from Ryckman et al. [17] and Ku et al. [18] to project which people with TB experience either death or spontaneous resolution before they receive any treatment.
2. For those who initiate treatment, we use WHO estimates of country-specific treatment outcomes to project the proportion of people who are cured through their initial treatment course [19]. For those in whom initial treatment does not result in lasting cure, we estimate a probability of eventual death versus eventual cure [17, 20-22].
3. Cumulative duration of sputum-positive disease, stratified by bacillary burden, is estimated by again drawing on projections from Ryckman et al. of the disease duration prior to any treatment among people without HIV, adjusting for HIV status, and extrapolating to estimate durations of recurrent TB disease after treatment failure or relapse.

Each of these steps is described in greater detail below. Of note, both Ryckman et al. and Ku et al. project TB disease outcomes and duration for people with symptomatic or asymptomatic TB disease, with only those with TB symptoms seeking care for TB under passive case-finding. Although there is variability in the criteria used for symptom screening and the reporting of symptoms between settings, we assumed that estimates for symptomatic and asymptomatic individuals, respectively, in Ryckman et al. and Ku et al. could be applied to individuals who did and did not report cough for > 2 weeks (“chronic cough”) in Uganda’s prevalence surey.

#### 3.1 Outcomes of TB episode prior to receiving treatment

Ryckman et al. fit a model simultaneously to historical TB survival data and to present-day TB prevalence, notification, and mortality data from several high-TB-, low-HIV-burden countries. Based on this data, Ryckman et al. first construct a hypothetical population of individuals with prevalent, symptom positive/negative and smear microscopy positive/negative TB at a certain point in time, giving equal weight to simulations from each of the five countries included in that analysis.

In a second step, assuming passive, symptom-based case finding to be in place, Ryckman et al. project the proportion of symptom positive/negative and smear positive/negative individuals who would eventually

1) receive treatment, or
2) experience spontaneous resolution prior to receiving treatment, or
3) experience death due to TB prior to receiving treatment.

In addition to the outcomes of TB disease, again for each proportion of prevalent, symptom positive/negative and smear positive/negative TB, Ryckman et al. estimate the average disease duration from the time of the cross-sectional sample until any of the TB disease outcomes are reached. This average disease duration is computed separately for time spent smear positive/negative and symptom positive/negative, considering that, for example, an individual might be smear- and symptom-positive at the time of the cross-sectional sample, but both smear and symptom status may change before any of the disease outcomes are reached.

While Ryckman et al. project outcomes and durations of TB only for HIV-negative people, our model requires estimates for HIV-positive people as well. We therefore adjust the TB outcomes and disease durations from Ryckman et al. for the effects of HIV infection on TB disease progression, mortality, and diagnosis, by drawing on a related modeling analysis of TB symptom onset and care-seeking behavior in several high- and low-HIV-burden settings by Ku et al.. For high-HIV-burden countries, Ku et al. used HIV-stratified prevalence, notification, and mortality estimates, together with assumptions about HIV and TB mortality rates, to estimate time from TB onset to diagnosis in each modeled country (Table S4 and S5). Our use of these results is described below.

##### 3.1.1 Estimating the proportion of people with prevalent TB experiencing death prior to receiving treatment, under routine care alone

For HIV-negative people (*h* = 0) with prevalent TB, our estimate of the proportion experiencing death prior to receiving treatment ($\mu_{h=0,b,s})$ is directly drawn from Ryckman et al. These proportions are stratified by each individual’s initial bacillary load at the time of a possible case-finding intervention: *b* = 1 for individuals with high bacillary load (defined as people with TB who are projected to test positive on smear microscopy in the Ryckman et al. model) and *b* = 0 for individuals with low bacillary load (i.e., those projected to test negative on smear microscopy by Ryckman et al.). The proportions are also stratified by initial presence of chronic cough (i.e., cough for > 2 weeks): *c* = 1 for individuals with chronic cough (defined as TB symptom positive in the Ryckman et al. model) and *c* = 0 for individuals without chronic cough .

For people with HIV (*h* = 1) and prevalent TB, we utilize the HIV-stratified prevalence-to-notification ratios and excess HIV-associated mortality estimated in Ku et al. to infer the proportion of HIV-positive people with prevalent TB who will experience death prior to TB treatment ($\mu_{h=1,b,c})$. TB duration is shorter in people with HIV, and faster disease progression results in both faster diagnosis and a higher rate of mortality prior to diagnosis. We estimate cumulative mortality by applying the estimated mortality rate over the estimated duration of undiagnosed TB. In detail, this approach comprises six steps:

1) For each initial bacillary load and chronic cough status in HIV-negative individuals, we start by drawing the time spent smear- and symptom- positive/negative until death due to TB, spontaneous resolution, or initiation of treatment from Ryckman et al’s model of TB disease trajectories in HIV-negative people. We assume that HIV status is unchanged for the duration of a TB episode, but the bacillary load and chronic cough status of individuals with TB can change over time in our model. Our notation distinguishes the initial smear and symptom status as b and c, and their time-varying counterparts as b’ and c’.
2) We adjust the durations $\delta_{h=0,b,c}$ drawn from Ryckman et al. to reflect TB shorter disease courses in people with HIV:

$$\delta_{h=1,b,c}=h_{duration}*\delta_{h=0,b,c}$$

   where
   - $h_{duration}$= adjustment of the TB disease duration for HIV positive people. Our estimate $h$ = 0.31 corresponds to the ratio of estimated disease durations in HIV-positive versus HIV-negative people in Kenya, in the version of Ku et al.’s model that assumed an additional monthly excess mortality rate of 5% for those people with TB who are HIV positive.

For a person with initial smear status *b* and initial presence of chronic cough *c,* the cumulative time $\delta_{h=1,b,c,b^{'},c^{'}}$that they spend in a given time-varying smear state (*b’*) and state of chronic cough (*c’*) is reduced by the same factor $h_{duration}$ if they are HIV-positive:

$$\delta_{h=1,b,c,b^{'},c^{'}}=h_{duration}*\delta_{h=0,b,c, b^{'},c^{'}}$$

3) Steps 3 to 5 estimate the relative probability of dying from TB prior to treatment in HIV positive compared to HIV negative people with TB. This is used to adjust $\mu_{h=0,b,c}$ in step 6. As a first step in computing this relative probability, we assume that average TB mortality rates in the overall population mirror the TB mortality rates prior to receiving treatment, and we estimate overall monthly mortality rates in people with TB, $\xi_{t=1,h,b^{'},c^{'}}$, as follows:

$$\xi_{t=1,h,b^{'},c^{'}}=\xi_{h}{+ \xi}_{t=1,h=0,b^{'},c^{'}}+\xi_{t=1,h}$$

where

- $\xi_{h}$ = a background mortality rate that applies to any individual:
  - estimated as 0.06% per month in HIV negative people ($\xi_{h=0})$, i.e., one-twelfth of the 2023 yearly crude death rate of 0.73% projected by the United Nations for Kenya [23]. We use United Nations estimates for Kenya to be internally consistent with our estimates of the HIV-dependence of disease duration which were based on Ku et al’s model of data from Kenya.
  - estimated as 0.11% per month in HIV positive people ($\xi_{h=1})$, to account for additional (non-TB-related) mortality associated with HIV infection. Using data from the UNAIDS AIDSinfo dashboard [24], we compute this mortality rate by
    - first dividing the number of HIV-related death in individuals without TB in Kenya in 2022 (estimated as the total number of ‘AIDS-related deaths – Adults (15+)’ [19,000] reduced by the total number of ‘TB-related deaths among people living with HIV’ [12,000]), by the total number of ‘People living with HIV’ in Kenya (1.3 million) and
    - adding this HIV-related mortality rate to $\xi_{h=0}$.

As above, we choose data from Kenya for internal consistency with estimates taken from Ku et al.

- $\xi_{t=1,h=0,b^{'},c^{'}}$ = an additional mortality rate in HIV-negative people with TB:
  - estimated as 0.5% per month during periods of low bacillary load-TB without chronic cough ($\xi_{t=1,h=0,b^{'}=0,c^{'}=1}$), based on the country-specific smear negative mortality rate estimates in the fitted model by Ryckman et al. and using the mean across all countries for consistency,
  - estimated as 3.0% per month during periods of high bacillary load TB with chronic cough ($\xi_{t=1,h=0,b^{'}=1,c^{'}=1})$, based on the country-specific smear positive mortality rate estimates in the fitted model by Ryckman et al. and using the mean across all countries for consistency
  - assumed to be 0% in TB without chornic cough of any bacillary load ($\xi_{t=1,h=0,b^{'},c^{'}=0}$).
- $\xi_{t=1,h,c^{'}=1}$= an additional mortality rate component reflecting increased mortality risks of HIV-associated TB with chronic chough, assumed equal to 5% per month if h=1 (for consistency with Ku et al.) and 0% if h=0.

4) We apply these monthly mortality rates $\xi_{t=1,h,b^{'},c^{'}}$ to our estimates of the pre-treatment disease duration $\delta_{h,b,s,b^{'},c^{'}}$ to compute a preliminary probability of death from TB prior to treatment (${\mu prelim}_{h,b,c}$):

${\mu prelim}_{h,b,c}$= $1- e^{-\sum_{b^{'},c^{'}} \xi_{t=1,h,b^{'},c^{'}}*\delta_{h,b,c,b^{'},c^{'}}}$

5) To ensure consistency with further values used from Ryckman et al., we do not directly use $\mu prelim$ in our model, but compute the ratio of these (τ) for HIV negative versus HIV positive people.

$$\tau=\frac{{\mu prelim}_{h=1,b,c}}{{\mu prelim}_{h=0,b,c}}$$

6) As a final step, we apply these ratios to the estimates of the proportion prevalent TB in HIV-negative people that will end in death prior to treatment under a routine care scenario ($\mu_{h=0,b,c})$from Ryckman et al. This provides our estimate of the probability of death prior to treatment for people with HIV and prevalent TB under a scenario of routine care only ($\mu_{h=1,b,c})$:

$$\mu_{h=1,b,s}=\tau*\mu_{h=0,b,c}$$

All of the parameters described above are listed in Table S4 and S5. In addition, for the ease of understanding, we provide an Excel file (“Additional file for Text S3.1.1 to S3.1.3.xlsx”) performing the above-described computations as well as those described in chapter 3.1.2 and 3.1.3 (below) using the point estimate of each described parameters.

##### 3.1.2 Proportion of people with prevalent TB who eventually receive TB treatment, under a scenario of routine care only

For HIV-negative people, similar to the approach above for estimating the probability of death (chapter 3.1.1), we take estimates from Ryckman et al. for the probability of eventual treatment under routine care, as a function of initial presence of chronic cough and initial bacillary burden. However, due to random sampling of all model parameters (chapter 4), in some instances, the cumulative proportion of those experiencing death prior to treatment and of those receiving treatment might exceed one. To avoid such situation, we apply these randomly sampled probabilities sequentially (instead of applying them as cumulative probabilities). Specifically, for the probability to receive treatment in HIV-negative people, we first compute the probability to eventually receive treatment relative to the cumulative probability of all outcomes but death prior to treatment in HIV-negative individuals ($\eta_{relative, h=0}$):

$$\eta_{relative, h=0}=\frac{\eta_{h=0}}{\eta_{h=0}+ \omega_{h=0}}$$

where

- $\eta_{h=0}$ = the probability to receive treatment for HIV-negative people as randomly sampled from Ryckman et al.
- $\omega_{h=0}$ = the propability to experience spontaneous resolution for HIV-negative people as randomly sampled from Ryckman et al.

Afterwards, we apply $\eta_{relative, h=0}$ to the proportion of HIV-negative individuals who were estimated not to experience death prior to receiving treatment $\left( 1-\mu_{h=0} \right)$:

$$\rho_{h=0}=\left( 1-\mu_{h=0} \right)* \eta_{relative, h=0}$$

where

- $\rho_{h=0}$ = the probability to receive treatment for HIV-negative people as eventually applied in our model.

For people with HIV, we estimate the probability of eventual treatment by assuming the relative probability of eventual treatment (HIV-positive versus HIV-negative) for prevalent TB in a cross-sectional sample to equal the ratio of TB case-detection ratios (i.e., the relative probability of eventual treatment of incident TB) of an HIV-positive versus HIV-negative incident TB case. Drawing on TB case-detection ratios estimated by Ku et al., we project the probability of eventual treatment to be higher for HIV-associated TB by a factor of $h_{treatment}=1.029$ (Table S5); this reflects the tendency of HIV-associated TB to progress more rapidly and consistently to a disease state with chronic cough and the resulting higher chance of being detected, as well as potentially more passive TB case-finding activity within the HIV care setting.

As above, to avoid any probabilities greater one, we use the probability of HIV-positive individuals with TB to receive treatment relative to all other outcomes but death prior treatment ($\eta_{relative, h=0}$). We adjust the odds of $\eta_{relative, h=0}$ for the impact of HIV-associated TB ($h_{treatment}$), converting the adjusted odds back to probabilities afterwards ($\eta_{relative, h=1})$. Lastly, we apply this adjusted probability to the proportion of individuals who were estimated not to experience death prior treatment, to project the proportion of HIV-positive individuals with TB receiving treatment ($\rho_{h=1}$):

$$\rho_{h=1}=\left( 1-\mu_{h=1} \right)* \eta_{relative, h=1}$$

##### 3.1.3 Proportion of people with prevalent TB experiencing spontaneous resolution without treatment, under routine care alone

We calculated the proportion of prevalent TB that will experience spontaneous resolution in the absence of being linked to treatment under routine care ($\Phi_{t,h,b,c}$) as:

$$\Phi_{h,b,c}=1-\mu_{h,b,c}-\rho_{h,b,c}$$

where

- $\mu_{h,b,c}$ = proportion of people with prevalent TB experiencing death without receiving treatment in a routine care setting (as above)
- $\rho_{h,b,c}$ = proportion of people with prevalent TB receiving treatment in a routine care setting.

The quantitative estimates can be found in Table S5 and in the main manuscript, Table 2.

#### 3.2 TB outcomes among those who start treatment

Drawing on country-specific, HIV-stratified treatment outcome estimates provided by WHO [19], we first estimated the proportions of treatment courses that end in (a) treatment failure, (b) loss to follow-up (LTFU) during treatment, and (c) death during treatment (Table S6); the remainder of treatment courses were classified programmatically as successful (with unknown treatment outcomes being excluded from this calculation). However, we modeled a possibility of cure among those LTFU and a risk of relapse among those considered to have been successfully treated, as follows.

Among treatments classified as programmatically successful, we assumed the risk of relapse to be 4% (95% uncertainty range: 3% to 10%). We based the point estimate of this uncertainty range on the relapse proportions estimated from treatment for drug susceptible TB in clinical trials, using pooled results from control arms that received standard regimens (4%; adjusted for the fact that only an estimated 75% of recurrence in the first two years after treatment occurs due to relapse) [20]. In the absence of a more accurate estimate, the lower bound was pragmatically defined as 3/4 of the point estimate. The upper bound was estimated using programmic outcomes of smear-positive drug-susceptible TB under India’s revised national tuberculosis control program during the era of thrice weekly treatment, were a multi-centric cohort study projected recurrence rates of 11%, >90% of which were due to relapse [22]. The remainder of programmatically successful treatments were assumed to result in cure.

Similarly, among treatments that ended in LTFU, we estimated the proportion who were cured by their partial treatment course, assuming that the timing of LTFU is distributed evenly across the six month treatment course [25] and that the proportion of people experiencing cure develops in a linear manner compared to the time between treatment initiation and LTFU [21] (Table S6).

We group the resulting estimates into three mutually exclusive outcomes of initial TB treatment:

1) Death
   a. All people who died during their initial TB treatment course
2) Cure
   a. Individuals who completed initial TB treatment successfully and did not relapse
   b. Individuals LTFU during initial treatment but who were cured by their partial treatment course
3) Failure/relapse
   a. All people in whom initial TB treatment failed
   b. Individuals in whose TB relapsed after initial treatment
   c. Individuals LTFU during initial treatment who were not cured

For those whose initial treatment ended in failure/relapse, we also model eventual outcomes after failure/relapse of either:

1) Eventual cure (through spontaneous resolution or one or more retreatments) or
2) Death

Considering that individuals receiving TB treatment after recurrence have poorer outcomes if diagnosed than those without prior TB disease [26] (and also likely a lower probability of spontaneous resolution), for all individuals who experience failure/relapse after initial treatment in our model, we assume an increased death probability equal to the probability of death before initial treatment for a person with chronic cough and high bacillary load TB (stratified by HIV status; Table S5). We model all remaining individuals to be eventually cured from TB.

#### 3.3 Estimating disease duration under routine care

To estimate the total time spent with prevalent TB in the target population, stratified by bacillary load as a proxy for transmission potential, we sum individual projections of cumulative bacillary-load-stratified disease durations (as a function of each case’s initial HIV, chronic cough, and bacillary load status) over all individuals with prevalent TB in the target population.

As a first step, we calculated a prevalence-weighted sum of average pre-treatment disease duration, in the absence of ACF, for all people with TB in the target population:

$$\delta_{total}= \sum_{h,b,{c,b}^{'},c'} \delta_{h,b,c, b^{'},c^{'}}*TB_{h, b,c}$$

where

$TB_{h, b,c}$ = the proportion of the target population that had TB with the specified initial HIV, bacillary load, and cough status.

We then repeat this calculation for b’=0 and, separately, for b’=1, to obtain the total cumulative treatment-naïve person-time that people with prevalent TB will spend with high and low bacillary load, respectively, in the absence of ACF:

$$\delta_{b^{'}=0}= \sum_{h,b,c,c'} \delta_{h,b,c, b^{'}=0,c^{'}}*TB_{h, b,c}$$

and

$$\delta_{b^{'}=1}= \sum_{h,b,c,c'} \delta_{h,b,c, b^{'}=1,c^{'}}*TB_{h, b,c} .$$

Afterwards, we account for additional disease duration that accrues after the first treatment attempt, for those who fail / relapse. We assume that the future cumulative duration of infectious TB after an initial failure / relapse, for people with a given HIV status, is the same as the duration of TB prior to any treatment. Thus, we again utilize the projections of pre-treatment TB disease duration from Ryckman et al. to estimate TB disease duration after the first treatment attempt. Because an individual’s cough and bacillary load status may differ from their initial status, these durations are an average of $\delta_{h,b,c, b^{'},c^{'}}$ over all b and c, weighted by the distribution of b and c in the target population, for a given h∈(0,1). Table S4 and S5).

### Text S4 – Methods Monte Carlo Simulation.

We performed a Monte Carlo simulation with 10,000 iterations. Each iteration represented a distinct cohort of 400,000 people.

For parameters that were bounded between zero and one and had clinical data that could be directly applied, we drew samples from a beta distribution (using the *rbeta* function from the *R base package*) with its mode at the source data’s point estimate and its beta value calculated based on confidence intervals or uncertainty ranges found in the literature. If a range was not reported in the literature, we estimated ranges from reported effect sizes and sample sizes using the *qbeta* function from the *R base package*. For parameters not bounded above, we fit a gamma distribution (using the *rgamma* function from the *R base package*), directly sampling from posterior distributions of model results provided by Ryckman et al. (Table S12 in the Appendix of Ryckmen et al.), or, where such was not available, drawing parameter ranges from 95% confidence intervals reported in the literature. For model-derived parameter estimates where uncertainty was greater, we drew parameter values from uniform distributions corresponding to the 95% uncertainty range reported by the prior modeling analysis (using the *runif* function from the *R base package*). For the TB disease durations in HIV-negative individuals derived from Ryckman et al., we directly draw 10,000 random samples from the posterior distributions of country-level simulations conducted in Ryckman et al. (to most closely mirror the original distribution of these parameters, see “Additional file random samples Ryckman et al”; the mean of the 10,000 random samples as reported in Figure 2 of Ryckman et al. is shown in Table S4).

Despite random sampling, we found that 10,000 model iterations were sufficient for stable results (Figure S6; in < 0.1% of the simulations, random sampling led to some of the proportions of the initial population falling below zero – respective simulations were dropped, resulting in 9,992 model iterations being included in the final analysis).

When a parameter estimate was changed in sensitivity or scenario analysis, the whole distribution was scaled accordingly (without changing the type of distribution). All computations were performed using R version 4.2.2. All parameters with literature-reported ranges / effect and sample sizes can be found in the main manuscript, Table 2, and in Tables S1 to S6, as well as in Table S7-8 for the sensitivity and scenario analysis.

### Text S5 – Methods for additional analyses.

#### 5.1 Minimal required sensitivity for oral-swab-based testing

Attention has been given recently to the potential to perform TB molecular testing on an oral swab instead of a sputum sample. Early data suggests that such an approach, although increasing the proportion of people from whom a sample can be collected, is likely to have lower sensitivity than sputum testing, potentially diminishing the benefits of its increased accessibility [27]. Therefore, in addition to our primary analyses in which test characteristics only improved, we explored potential trade-offs between specimen availability (with a non-sputum test) and test sensitivity. We estimated the minimal sensitivity required for oral-swab based molecular test (assuming the same cost and turn-around time as sputum-based molecular testing, but an increase in specimen collection from 93% to 100%) to achieve at least the same reductions in TB mortality and transmission as sputum Xpert Ultra testing, when used for confirmatory testing in ACF.

We estimated the number of incremental TB-related deaths averted and the incremental transmission potential averted, compared to routine care alone, through ACF with oral-swab based confirmatory testing at different sensitivity levels. We considered sensitivity levels for oral-swab based testing of 0%, 15%, 30%, 70%, 80%, 90%, and 100% relative to sputum-based confirmatory testing. For the relative sensitivities of 70%, 80%, and 90%, we assumed any reduction in test sensitivity to result from reduced detection of individuals with low bacillary load [28]. For the relative sensitivities of 15% and 30%, we assumed that none of the individuals with low bacillary load would be detected and any further sensitivity reductions to result from decreased detection of individuals with high bacillary load. Afterwards, we fit a quadratic regression line (using the R function *ggplot_smooth*), with the slope equaling the incremental number of TB deaths averted and the incremental TB transmission potential averted per increase in confirmatory test sensitivity. Based on this line, we projected the test sensitivity at which oral-swab based confirmatory testing would result in at least as many TB deaths and as much TB transmission potential averted as sputum-based confirmatory testing. To account for uncertainty in the proportion of individuals that are able to produce sputum, we performed this analysis assuming 93% (as in the primary analysis), 83% and 73% of individuals able to produce sputum.

#### 5.2 One-way sensitivity analysis of country-specific parameters

To explore the dependence of our results on epidemiological characteristics of the model setting, we performed one-way sensitivity analyses that individually varied prevalence of TB and TB/HIV co-infection, as well as chest X-ray and non-cartridge costs of the confirmatory test.

As upper ranges for the sensitivity analysis, mirroring a South-Africa-like setting, we assumed a TB prevalence of 3,408 / 100,000 (assuming, as in the baseline analysis, a TB prevalence *four* times the national prevalence) [3]. To further explore the impact of changes in the TB epidemic on our model results, in another analysis, we assumed 74.3% of prevalent TB to have a high bacillary load (using data from Nigeria) [29]. Considering the influence of HIV on TB disease outcomes in our model, we further assumed an increased HIV prevalence of 17,070 / 100,000 [3]. Reflecting potential changes in test costs, we also modelled an increase in the non-cartridge costs of the confirmatory test by ca. 87% (projecting labor costs for chest X-ray screening to increase to the same extent) [30].

The lower ranges of our one-way sensitivity analysis reflected an India-like setting. Moreover, to test our model’s robustness to more extreme ranges, we assumed community-based screening in India would be conducted in a setting with a TB prevalence only *twice* the national prevalence – in contrast to the high TB-prevalence setting of *four* times Uganda’s national prevalence assumed in our baseline analysis. Based on these assumptions, in a first analysis, we modeled a TB prevalence of 632 / 100,000 [31]. In a second analysis, we estimated 32.8% of prevalent TB to have a high bacillary load (using data from Cambodia) [32]. We also considered an HIV-prevalence of 23 / 100,000 [33], as well as a decrease in Xpert Ultra non-cartridge costs by ca. 38% [34] (estimating labor costs for chest X-ray screening to decrease to the same extent) (Table S7).

#### 5.3 Scenario-analyses of further underlying model assumptions

We assessed the extent to which the benefit of different confirmatory test improvements changed when the screening test was changed from using chest X-ray to screening for the presence of chronic cough, when treatment costs were included in the ACF budget, or when the primary model’s high levels of spontaneous TB resolution were reduced.

When modeling symptom screening (i.e., for chronic cough) in place of X-ray, the costs of screening were estimated at $2.21 per person screened [35]. For the scenario analysis in which treatment costs were part of the active case-finding (ACF) budget, we included increased confirmatory test specificity as one of the test improvements considered.

Given uncertainty around the proportion of prevalent TB experiencing spontaneous resolution, we reevaluated our results for TB deaths averted if the proportion of TB spontaneously resolving were reduced by 50% and all outcomes were distributed between death before treatment and treatment initiation according to their relative proportions. We also modeled the effect of confirmatory test improvement on TB deaths averted if no TB spontaneously resolved. We did not consider changes in transmission for these scenarios, because the disease durations drawn from Ryckman et al. (Text S3) were no longer valid after eliminating spontaneous resolution.

### Text S6 – Results of the sensitivity and scenario analysis

#### 6.1 – Results of the sensitivity analysis

We performed one-way sensitivity analyses, separately varying the TB prevalence, the proportion of TB that had high bacillary load, the prevalence of TB/HIV co-infection, and test costs. We found that a decrease in the proportion of prevalent TB with high bacillary load (from the baseline estimate of 41% to 33% of high-bacillary-TB), resulted in a larger estimate of the impact of increased test sensitivity (from 69% to 80%) on TB mortality and transmission potential: the number of TB deaths averted by ACF increased from 5% (2-11%) in the primary analysis to 7% (2-15%) when the proportion of individuals with high TB bacillary load was lower, and the transmission potential averted increased from 5% (2-8%) to 6% (2-11%) (Figure S2).

Another noteworthy difference was seen if non-cartridge costs were higher (assuming non-cartridge costs of $23 instead of $12 per confirmatory test). In such as setting, the impact of reducing confirmatory test costs (by the same 50% factor as in the primary analysis) lead to an increase in the impact on TB mortality and transmission potential relative to ACF with the baseline confirmatory test from 6% (2-22%) to 9% (3-30%) (Figure S2). Also, in this sensitivity analysis the relative importance of the confirmatory test improvements changed, with reducing test costs becoming the test improvement with the second greatest impact on TB mortality and transmission potential instead of using a non-sputum specimen as in the primary analysis (Figure S2).

#### 6.2 – Results of the scenario analysis assuming that a tuberculosis infection cannot spontaneously resolve

When we reduced the possibility of spontaneous TB resolution by 50%, under routine care alone, 1,263 (920-1,670) of the 6,421 (5,316-7,531) individuals with TB were estimated to die from TB. Under the allocated budget of one million USD, 83,808 individuals (59,388-118,601) could be included in ACF efforts using the baseline confirmatory test. Compared to routine care alone, this would avert 93 (51-159) deaths, resulting in total of 1,166 (859-1,535) individuals projected to die from TB.

Relative to ACF using the baseline confirmatory test, greatest reductions in TB mortality were projected for immediate turn-around of test results (11% [5-18%] more deaths averted; 10 [4 to 20] death averted) and increasing test sensitivity (10% [3-21%] more deaths averted than through ACF using the baseline confirmatory test). This was followed by using a non-sputum specimen (8% [4-12%] more deaths averted) and reduced test costs (6% [2-22%] more deaths averted) (Figure S3).

Assuming no TB to spontaneously resolve, 1,645 (1,196-2,211) of all 6,421 (5,316-7,531) prevalent TB was projected to die. With 83,808 individuals (59,388-118,601; as under reduced spontaneous resolution) possible to include in ACF under the allotted budget, ACF could avert 111 (61-188) TB deaths. Improving confirmatory testing largely resulted in the same benefit on TB mortality as with reduced spontaneous resolution (above). Only increased test sensitivity lead to a larger effect, reducing TB mortality by 13% (4-28%; 14 [4-33] incremental deaths averted) relative to ACF using the baseline confirmatory test.

Potential changes in transmission potential were not considered for these scenarios, since, after assuming that TB cannot spontaneously resolve, the disease durations used from Ryckman et al. (Text S3) were no longer applicable (Text S5).

#### 6.3 – Results of the scenario analysis including treatment costs as part of the active case-finding budget

To evaluate the potential benefit of increased confirmatory test specificity, we also considered treatment costs as part of the ACF budget. Under this scenario, relative to the baseline confirmatory test, an increase in test specificity (from 99% to 100%) increased the incremental deaths and transmission potential averted by active case-finding relative to routine care alone from 0% (0-0%) to 1% (0-3%). However, this was only a fraction of the potential impact of the next least impactful test improvement (immediate turn-around of test results; 10% [5-17%]) and did not affect the relevance of any of the other test improvements (Figure S4).

### Table S1 – Parameters describing the initial state of the model’s population.

| **Parameter** | **Point estimate** | **Lower range** | **Upper range** | **Assumed distribution** | **Source** |
| --- | --- | --- | --- | --- | --- |
| Proportion of ACF target population that have bacteriologically confirmed TB disease (four times the national prevalence) | 1.6% | 1. 2% | 2.0% | Beta | [1] |
| Proportion of ACF target population that are infected with HIV | 5.8% | 5.3% | 6.3% | Beta | [36] |
| Proportion of ACF target population that have cough > 2 weeks | 6.5% | 1.2% * | 15.7% * | Beta | [1] |
| Proportion of ACF target population with cough > 2 weeks that are infected with HIV | 28.6% | 22.6% | 35. 5% | Beta | [33] |
| Proportion of ACF target population with bacteriologically confirmed TB that have a high TB bacillary load | 41.3% | 33.8% * | 48.9% * | Beta | [1] |
| Proportion of ACF target population with bacteriologically confirmed TB that are also infected with HIV ^a^ | 37.4% | 26.9% | 48.0% | Beta | [1, 37] |
| Proportion of ACF target population with bacteriologically confirmed TB that also have cough > 2 weeks | 49.4% | 41.7% * | 57.1% * | Beta | [1] |
| Proportion of ACF target population with bacteriologically confirmed TB that have cough > 2 weeks and are HIV infected | 33.3% | 19.6% * | 48.7% * | Beta | [33] |
| Proportion of ACF target population with bacteriologically confirmed TB and HIV infection that have a high TB bacillary load | 30.8% | 9.9% * | 57.2% * | Beta | [33] |
| Proportion of ACF target population with bacteriologically confirmed TB and cough > 2 weeks that have a high TB bacillary load | 45.6% | 34.8% * | 56.6% * | Beta | [1] |
| Proportion of ACF target population with bacteriologically confirmed TB, HIV infection and cough > 2 weeks that have a high TB bacillary load | 30.8% | 9.9% * | 57.2% * | Beta | [33] |

^a^ The proportion of bacteriologically confirmed TB that are also infected with HIV is reported as 26.9% in a Uganda prevalence survey [1]. However, authors of this survey highlight that this value is much lower than generally accepted WHO estimates for Uganda, reporting 48.0% of prevalent TB being HIV co-infected [37]. To account for the uncertainty in this estimate, we chose the mean of both reports as the point estimate (37.4%), with the other two values as the lower and upper range, respectively.

* Lower and upper range were estimated based on the parameters’ point estimate and sample size (as provided in the literature), using the *qbeta* function from the R base packages

### Table S2 – Parameters to estimate the accuracy of diagnostic tools.

| **Test** | **Parameter** | **Reference population** | **Point estimate** | **Lower range** | **Upper range** | **Assumed distribution** | **Source** |
| --- | --- | --- | --- | --- | --- | --- | --- |
| CXR | Sensitivity | Average across the total population | 90.0% | 84.9% * | 94.1% * | Beta | [1] |
| CXR | Sensitivity | Only in people with TB that have a high bacillary load | 100% | 100% | 100% | Beta | [1] |
| CXR | Specificity | Average across the total population | 96.0% | 93.0% | 97.0% | Beta | [2] |
| CXR | Specificity | Only in people with cough > 2 weeks | 81.3% | 79.8% * | 82.8% * | Beta | [1] |
| CXR | Specificity | Only in people with HIV infection | 87.2% | 86.2% * | 88.1% * | Beta | [3] |
| Xpert | Sensitivity | Average across the total population | 69.0% | 48.0% | 86.0% | Beta | [2] |
| Xpert | Sensitivity | Only in people with TB that have a high bacillary load | 99.1% | 97.8% * | 99.8% * | Beta | [4] |
| Xpert | Specificity | Average across the total population | 98.8% | 97.2% | 99.5% | Beta | [2] |

* lower and upper range were estimated based on the parameter’s point estimate and sample size (as provided in the literature), using the qbeta function from the R base packages

### Table S3 – Cost parameters for TB testing and treatment.

| **Category** | **Point estimate** | **Lower**  **range** | **Upper**  **range** | **Assumed distribution** | **Source** |
| --- | --- | --- | --- | --- | --- |
| Equipment costs for screening (for two years; 2023 USD) ^a^ | 75,018.00 | 56,263.50 | 93,772.50 | Gamma | [8, 9] |
| Staff costs for screening (for two years; 2023 USD) ^a^ | 177,170.15 | 132,877.61 | 221,462.68 | Gamma | [12] |
| Confirmatory test cartridge costs (per test; 2023 USD) ^b^ | 7.97 | 7.97 | 7.97 | Gamma | [13] |
| Confirmatory test labor/equipment costs (per test; 2023 USD) ^b^ | 12.09 | 9.08 | 18.15 | Gamma | [14] |
| Treatment costs (per treatment; 2023 USD) | 159.42 | 75.63 | 243.21 | Gamma | [15] |
| Number of people that can be screened with one set of chest X-ray equipment and staff within two years | 25,000 | 18,750 | 31,250 | Gamma | Text S3 |

^a^ Assuming chest X-ray as the screening tool

^b^ Assuming sputum-based Xpert Ultra as the confirmatory test

### Table S4 – Estimated untreated TB disease durations under routine care for HIV-negative individuals, drawn from simulations by Ryckman et al [17]

| **Initial TB state** | | **TB state passed during TB disease course** | **Time spent in each TB state** (mean number of months [2.5^th^ and 97.5^th^ percentile])^c^ |
| --- | --- | --- | --- |
| **Smear status**^a^ | **Symptoms**^b^ |  |  |
| Low | Negative | Smear negative, asymptomatic | 2.5 [1.8−4.7] |
| Low | Negative | Smear positive, asymptomatic | 0.9 [0.4−1.5] |
| Low | Negative | Smear positive, symptomatic | 0.8 [0.4−1.7] |
| Low | Negative | Smear positive, symptomatic | 0.7 [0.3−1.1] |
|  | | **Total time with (any) TB** | 4.8 [3.3−8.4] |
| High | Negative | Smear negative, asymptomatic | 0.3 [0.0−0.7] |
| High | Negative | Smear positive, asymptomatic | 9.6 [5.9−15.3] |
| High | Negative | Smear positive, symptomatic | 0.1 [0.0−0.3] |
| High | Negative | Smear positive, symptomatic | 5.9 [3.7−8.9] |
|  | | **Total time with (any) TB** | 15.9 [11.1−23.4] |
| Low | Positive | Smear negative, asymptomatic | 2.0 [1.1−3.8] |
| Low | Positive | Smear positive, asymptomatic | 1.2 [0.5−2.0] |
| Low | Positive | Smear positive, symptomatic | 3.1 [2.1−5.1] |
| Low | Positive | Smear positive, symptomatic | 1.3 [0.7−2.0] |
|  | | **Total time with (any) TB** | 7.6 [5.3−10.9] |
| High | Positive | Smear negative, asymptomatic | 0.2 [0.0−0.5] |
| High | Positive | Smear positive, asymptomatic | 4.4 [0.3−8.8] |
| High | Positive | Smear positive, symptomatic | 0.1 [0.0−0.3] |
| High | Positive | Smear positive, symptomatic | 6.3 [3.9−9.7] |
|  | | **Total time with (any) TB** | 11.0 [6.2−16.0] |
| Any | Any | Smear positive, symptomatic | 1.7 [1.1−3.2] |
| Any | Any | Smear positive, symptomatic | 3.0 [1.4−4.5] |
| Any | Any | Smear positive, symptomatic | 1.0 [0.5−2.0] |
| Any | Any | Smear positive, symptomatic | 2.5 [1.4−4.1] |
|  | | **Total time with (any) TB** | 8.2 [5.9−10.9] |

^a^ Classified as high/low bacillary load in our model.

^b^ Assumed to correspond in our model to indivivduals with/without chronic cough (i.e., cough for > 2 weeks), as this was the symptom criterion used in Uganda’s national prevalence survey that was used to characterize our modeled target population.

^c^ In our model, we draw 10,000 random samples of the disease durations based on the posterior distributions of the individual-level microsimulations conducted in Ryckman et al.. For the ease of notation, this table presents the mean and 2.5th and 97.5th percentiles of these random samples as shown in Figure 2 of Ryckman et al.

### Table S5 – Parameters used to estimate TB disease outcomes under routine care for people with HIV.

| **Parameter** | | **Point estimate** | **Lower range** | **Upper range** | **Assumed distribution** | **Source** |
| --- | --- | --- | --- | --- | --- | --- |
| Baseline TB disease outcomes of HIV-negative individuals ^a^ | | | | | | |
| Proportion of HIV-negative prevalent TB that will end in death without treatment under a routine care scenario ^b^ | Symptomatic, smear-positive | 75% | 53% | 90% | Uniform | [17] |
|  | Symptomatic, smear-negative | 27% | 15% | 50% | Uniform | [17] |
|  | Asymptomatic, smear-positive | 71% | 49% | 87% | Uniform | [17] |
|  | Asymptomatic, smear-negative | 10% | 6% | 18% | Uniform | [17] |
| Proportion of HIV-negative prevalent TB that will end in treatment under a routine care scenario ^b^ | Symptomatic, smear-positive | 18% | 8% | 30% | Uniform | [17] |
|  | Symptomatic, smear-negative | 5% | 2% | 9% | Uniform | [17] |
|  | Asymptomatic, smear-positive | 17% | 8% | 28% | Uniform | [17] |
|  | Asymptomatic, smear-negative | 2% | 1% | 4% | Uniform | [17] |
| Parameters to adjust the TB disease outcomes of HIV-negative individuals (above) for the impact of HIV infection | | | | | | |
| TB case detection ratio in HIV positive people | | 71% | 63% | 80% | Uniform | [18] |
| TB case detection ratio in HIV negative people | | 69% | 59% | 81% | Uniform | [18] |
| Background mortality rate (yearly) | | 0.78% | 0.70% | 0.81% | Beta | [23] |
| Additional mortality in HIV-negative people with  smear-negative TB (monthly) ^c^ | | 1% | 0% | 1% | n/a | [17] |
| Additional mortality in HIV-negative people with  smear-positive TB (monthly) ^c^ | | 3% | 2% | 5% | n/a | [17] |
| Excess mortality rate for HIV positive people with TB (monthly) | | 5% | 5% | 5% | Beta | [18] |
| Total TB disease duration in HIV negative people (months) | | 6.49 | 4.94 | 8.11 | Gamma | [18] |
| Total TB disease duration in HIV positive people (months) | | 20.90 | 18.01 | 24.46 | Gamma | [18] |
| People living with HIV, Kenya (number of people) | | 1,300,000 | 1,200,000 | 1,600,000 | Gamma | [24] |
| HIV related deaths, Kenya (yearly; number of deaths) | | 19,000 | 14,000 | 31,000 | Gamma | [24] |
| Deaths related to HIV/TB co-infection, Kenya (yearly; number of deaths) | | 12,000 | 6,200 | 19,000 | Gamma | [24] |

* Lower and upper range were estimated based on the parameters’ point estimate and sample size (as provided in the literature), using the *qbeta* function from the R base packages

^a^ All individuals that under routine care neither receive treatment nor experience death prior TB treatment, are modeled to spontaneously resolve prior to receiving any TB treatment.

^b^ Using the raw outputs of the model from Ryckman et al. [17]. For the purpose of our model, we define those individuals considered smear-microscopy positive/negative in Ryckman et al. as indivivduals with high/low TB bacillary load and those individuals considered symptom positive/negative in Ryckman et al. as indivivduals with/without chronic cough (i.e., cough for > 2 weeks) (Text S3).

^c^ In our model, we draw 10,000 random samples of the additional monthly mortality in HIV-negative individuals with smear-positive and -negative TB based on the posterior distributions of the individual-level microsimulations conducted in Ryckman et al.. For the ease of notation, this table presents the mean and 2.5th and 97.5th percentiles of these random samples based on Table S6 in the Appendix of Ryckman et al.

### Table S6 – Parameters to estimate TB treatment outcomes.

| **Category** | **Point estimate** | **Lower range** | **Upper range** | **Assumed distribution** | **Source** |
| --- | --- | --- | --- | --- | --- |
| Treatment failure in HIV negative people | 0.7% | 0.6% * | 0.8% * | Beta | [19] |
| Death during treatment in HIV negative people | 6.0% | 5.8% * | 6.3% * | Beta | [19] |
| Loss to follow up during treatment in HIV negative people | 9.1% | 8.8% * | 9.3% * | Beta | [19] |
| Treatment failure in HIV positive people | 0.5% | 0.4% * | 0.6% * | Beta | [19] |
| Death during treatment in HIV positive people | 11.7% | 11.1% * | 12.1% * | Beta | [19] |
| Loss to follow up during treatment in HIV positive people | 7.1% | 6.8% * | 7.5% * | Beta | [19] |
| Relapse (i.e., failure to cure) after treatment completion | 5% | 3% | 10% | Beta | [20, 22] |
| Relapse among those lost to follow up during treatment | 50% | 35% | 65% | Uniform | [21] |

* Lower and upper range were estimated based on the parameter’s point estimate and sample size (as provided in the literature), using the *qbeta* function from the R base packages

### Table S7 – Parameter ranges explored in one-way Sensitivity Analysis.

| **Parameter** | **Baseline Estimate**  **(lower – upper range)** | **Source** | **Lower Estimate**  **(lower – upper range)** | **Source** | **Upper Estimate**  **(lower – upper range)** | **Source** |
| --- | --- | --- | --- | --- | --- | --- |
| Prevalence  of TB | 1.6%  (1.2% to 2.0%) | Table S1 | 0.63%  (0.58% to 0.68%) ^a^ | [31] | 3.41%  (2.72% to 4.10%) | [3] |
| Proportion prevalent TB with high bacillary load | 41.3%  (36.7%* to 45.9%*) | Table S1 | 32.8%  (27.7%* to 38.1%*) | [32] | 74.3%  (66.9%* to 88.1%*) | [29] |
| Prevalence  of HIV | 5.8%  (5.3% to 6.3%) | Table S1 | 0.21%  (0.17% to 0.25%) | [38] | 17.07%  (16.62% to 17.52%) | [3] |
| Labor costs for screening (per screening test performed) | $6.02  ($4.13 to $7.15) | Table S5 | $4.38  ($3.29 to $5.48) ^c^ | [34] | $12.22  ($9.17 to $15.28) ^c^ | [30] |
| Labor/equipment costs of the confirmatory test (per confirmatory test performed) | $12.09  ($9.08 to $18.15) | Table S5 | $7.50  ($5.62 to $9.37) | [34] | $22.58  ($20.28 to $24.88) | [30] |

^a^ In the baseline analysis, we consider ACF to be conducted in a *high TB-prevalence* -- i.e., four times the national prevalence -- Uganda-like setting. To test our model’s behavior to a more extreme extent, for the lower estimate of the TB prevalence sensitivity-analysis, we consider active-case finding to be conducted in a *lower TB-prevalence* -- i.e., two times the national prevalence of TB -- India-like setting.

^c^ The lower and upper estimate of the labor costs for screening were computed adjusting the baseline estimate by the same ratio as between the lower and upper estimate of non-cartridge costs for confirmatory testing.

* Lower and upper range were estimated based on the parameter’s point estimate and sample size (as provided in the literature), using the *qbeta* function from the R base packages

### Table S8 – Parameters Scenario Analysis.

| Scenario | Parameter | Baseline Estimate | Source | Scenario Estimate | Source |
| --- | --- | --- | --- | --- | --- |
| No spontaneous resolution of TB | Under the baseline scenario, we estimate the proportion of prevalent TB that will end in spontaneous resolution under a routine care scenario (φ) as:  φ = 1 - proportion of prevalent TB that will end in death under a routine care scenario (∂) - proportion of prevalent TB that will end in cure under a routine care scenario (µ) (further details in Text S3 and Table S5).  To reduce the proportion of prevalent TB that spontaneously resolves under a routine care scenario to zero, after sampling for the uncertainty ranges, we compute ∂_adjusted_ and µ_adjusted_ as:  ∂_adjusted_ = ∂ / (∂ + µ) and µ_adjusted_ = µ / (∂ + µ). | | | | |
| Symptom screening^a^ | Sensitivity of chronic cough screening in people with symptomatic TB | n/a | n/a | 1.00 (1.00 to 1.00) | n/a |
|  | Sensitivity of symptom screening in people with asymptomatic TB ^a^ | n/a | n/a | 0.00 (0.00 to 0.00) | n/a |
|  | Specificity of symptom screening in people with symptomatic TB | n/a | n/a | 0.00 (0.00 to 0.00) | n/a |
|  | Specificity of symptom screening in people with asymptomatic TB | n/a | n/a | 1.00 (1.00 to 1.00) | n/a |
|  | Screening costs (per person screened) | $10 ($7 to $14) | Text S3 | $2.21 ($1.66 to $2.76) | [35] |

^a^ Using cough > 2 weeks to differentiate between people screening positive (i.e., having cough > 2 weeks) and screening negative.

### Table S9 – Population outcomes under active case-finding using Xpert Ultra compared to routine care alone.

| Population | Number of people under routine care alone  [95% UR] | Number of people under ACF using  Xpert Ultra  [95% UR] | Absolute change through ACF (Xpert Ultra) compared to routine care [95% UR] | Relative change through ACF (Xpert Ultra) compared to routine care [95% UR] |
| --- | --- | --- | --- | --- |
| Total Population | 400,000 | 400,000 | 0 [0; 0] | 0% [0%; 0%] |
| All people with TB started on treatment | 2716 [2089; 3420] | 3025 [2372; 3749] | 297 [181; 480] | 11% [6%; 19%] |
| >> All people with high bacillary load TB started on treatment | 1910 [1471; 2425] | 2025 [1580; 2541] | 113 [60; 192] | 6% [3%; 11%] |
| >> All people with TB and present chronic cough started on treatment | 1621 [1180; 2135] | 1758 [1316; 2276] | 132 [75; 221] | 8% [4%; 15%] |
| All people with TB cured | 2466 [1897; 3105] | 2766 [2175; 3424] | 289 [177; 468] | 12% [7%; 20%] |
| >> All people with high bacillary load TB cured | 1737 [1337; 2210] | 1847 [1439; 2317] | 107 [56; 181] | 6% [3%; 11%] |
| >> All people with TB and present chronic cough cured | 1468 [1069; 1934] | 1600 [1202; 2068] | 127 [73; 214] | 9% [5%; 16%] |
| All people with TB who die due to TB | 873 [612; 1182] | 796 [565; 1069] | -76 [-132; -39] | -9% [-13%; -6%] |
| >> All people with high bacillary load TB who die due TB | 638 [414; 903] | 570 [378; 802] | -67 [-118; -32] | -10% [-15%; -7%] |
| >> All people with TB and present chronic cough who die due TB | 516 [330; 751] | 472 [306; 680] | -43 [-83; -19] | -8% [-13%; -5%] |
| All people with TB that spontaneously resolve | 3047 [2384; 3790] | 2829 [2206; 3521] | -212 [-355; -126] | -7% [-11%; -4%] |
| >> All people with high bacillary load TB that spontaneously resolve | 254 [73; 472] | 212 [61; 399] | -39 [-82; -11] | -16% [-22%; -11%] |
| >> All people with TB and present chronic cough that spontaneously resolve | 1156 [796; 1611] | 1070 [738; 1487] | -83 [-147; -44] | -7% [-11%; -4%] |

### Table S10 – Population outcomes under community-based screening using Xpert Ultra or a hypothetical improved test for confirmatory testing.

| **Population Parameter** | Benefit of ACF (using Xpert Ultra) compared to routine care [95% UR] | Additional benefit through increasing the sensitivity of confirmatory testing | |
| --- | --- | --- | --- |
|  |  | Absolute  [95% UR] | Relative  [95% UR] |
| Total population | 400,000 | 0 [0; 0] | 0% [0%; 0%] |
| Included in ACF efforts | 83,808 [59,388; 118,601] | 0 [0; 0] | 0% [0%; 0%] |
| All people with TB started on treatment under ACF | 870 [578; 1,294] | 93 [29; 180] | 14% [4%; 26%] |
| >> All people with high bacillary load TB started on treatment | 515 [342; 776] | 4 [2; 8] | 1% [0%; 2%] |
| >> All people with TB and chronic cough started on treatment | 473 [312; 719] | 43 [14; 85] | 13% [4%; 23%] |
| All people with TB started on treatment under ACF that would have *died* in the absence of ACF | 83 [43; 145] | 4 [1; 10] | 5% [2%; 11%] |
| >> All people with high bacillary load TB started on treatment under ACF that would have *died* in the absence of ACF | 73 [36; 130] | 1 [0; 2] | 1% [0%; 2%] |
| >> All people with TB and chronic cough started on treatment under ACF that would have *died* in the absence of ACF | 47 [21; 91] | 3 [1; 7] | 6% [2%; 16%] |
| All people with TB started on treatment under ACF that would have *spontaneously resolved* in the absence of ACF | 212 [126; 355] | 66 [19; 131] | 31% [8%; 63%] |
| >> All people with high bacillary load TB started on treatment under ACF that would have *spontaneously resolved* in the absence of ACF | 39 [11; 82] | 0 [0; 1] | 1% [0%; 2%] |
| >> All people with TB and chronic cough started on treatment under ACF that would have *spontaneously resolved* in the absence of ACF | 83 [44; 147] | 25 [7; 52] | 31% [8%; 64%] |
| All people with TB started on treatment under ACF that would have *received treatment* in the absence of ACF | 351 [227; 540] | 22 [8; 45] | 6% [2%; 12%] |
| All people with TB started on treatment under ACF that would have *received treatment, but eventually died* in the absence of ACF | 32 [20; 50] | 2 [1; 4] | 7% [2%; 12%] |
| All people with TB started on treatment under ACF that would have *received treatment and eventually be cured* in the absence of ACF | 319 [207; 491] | 19 [7; 41] | 6% [2%; 11%] |
| All people with TB eventually cured following ACF | 611 [403; 924] | 90 [28; 175] | 15% [5%; 27%] |
| >> All people with high bacillary load TB eventually cured | 381 [252; 576] | 4 [1; 8] | 1% [0%; 2%] |
| >> All people with TB and chronic cough eventually cured | 315 [205; 483] | 41 [13; 81] | 13% [4%; 24%] |
| All people with TB cured on the first treatment attempt following ACF | 579 [381; 875] | 88 [27; 171] | 15% [5%; 28%] |
| >> All people with high bacillary load TB cured on the first treatment attempt following ACF | 352 [233; 533] | 3 [1; 7] | 1% [0%; 2%] |
| >> All people with TB and chronic cough cured on the first treatment attempt following ACF | 296 [193; 455] | 40 [13; 79] | 14% [4%; 25%] |
| All people with TB eventually cured following ACF that would have *died* in the absence of ACF | 76 [39; 132] | 4 [1; 9] | 5% [2%; 11%] |
| >> All people with high bacillary load TB eventually cured following ACF that would have *died* in the absence of ACF | 67 [32; 118] | 1 [0; 2] | 1% [0%; 2%] |
| >> All people with TB and chronic cough eventually cured following ACF that would have *died* in the absence of ACF | 43 [19; 83] | 2 [1; 6] | 6% [2%; 15%] |
| All people with TB who die due to TB | -76 [-132; -39] | -4 [-9; -1] | 5% [2%; 11%] |
| >> All people with high bacillary load TB who die due TB | -67 [-118; -32] | -1 [-2; 0] | 1% [0%; 2%] |
| >> All people with TB and chronic cough who die due TB | -43 [-83; -19] | -2 [-6; -1] | 6% [2%; 15%] |
| All people with TB that spontaneously resolve prior to receiving any treatment | -212 [-355; -126] | -66 [-131; -19] | 31% [8%; 63%] |
| >> All people with high bacillary load TB that spontaneously resolve prior to receiving any treatment | -39 [-82; -11] | 0 [-1; 0] | 1% [0%; 2%] |
| >> All people with TB and chronic cough that spontaneously resolve prior to receiving any treatment | -83 [-147; -44] | -25 [-52; -7] | 31% [8%; 64%] |

| **Population Parameter** | Benefit of ACF (using Xpert Ultra) compared to routine care [95% UR] | Additional benefit through conducting confirmatory testing on a non-sputum specimen | |
| --- | --- | --- | --- |
|  |  | Absolute  [95% UR] | Relative  [95% UR] |
| Total population | 400,000 | 0 [0; 0] | 0% [0%; 0%] |
| Included in ACF efforts | 83,808 [59,388; 118,601] | 0 [0; 0] | 0% [0%; 0%] |
| All people with TB started on treatment under ACF | 870 [578; 1,294] | 49 [26; 89] | 8% [5%; 12%] |
| >> All people with high bacillary load TB started on treatment | 515 [342; 776] | 31 [16; 56] | 8% [4%; 12%] |
| >> All people with TB and chronic cough started on treatment | 473 [312; 719] | 20 [7; 45] | 6% [2%; 12%] |
| All people with TB started on treatment under ACF that would have *died* in the absence of ACF | 83 [43; 145] | 6 [3; 13] | 7% [4%; 12%] |
| >> All people with high bacillary load TB started on treatment under ACF that would have *died* in the absence of ACF | 73 [36; 130] | 5 [2; 11] | 8% [4%; 12%] |
| >> All people with TB and chronic cough started on treatment under ACF that would have *died* in the absence of ACF | 47 [21; 91] | 3 [1; 7] | 6% [2%; 12%] |
| All people with TB started on treatment under ACF that would have *spontaneously resolved* in the absence of ACF | 212 [126; 355] | 17 [8; 33] | 8% [5%; 12%] |
| >> All people with high bacillary load TB started on treatment under ACF that would have *spontaneously resolved* in the absence of ACF | 39 [11; 82] | 3 [1; 7] | 8% [4%; 13%] |
| >> All people with TB and chronic cough started on treatment under ACF that would have *spontaneously resolved* in the absence of ACF | 83 [44; 147] | 5 [2; 12] | 6% [2%; 12%] |
| All people with TB started on treatment under ACF that would have *received treatment* in the absence of ACF | 351 [227; 540] | 26 [13; 48] | 7% [4%; 12%] |
| All people with TB started on treatment under ACF that would have *received treatment, but eventually died* in the absence of ACF | 32 [20; 50] | 2 [1; 4] | 7% [4%; 12%] |
| All people with TB started on treatment under ACF that would have *received treatment and eventually be cured* in the absence of ACF | 319 [207; 491] | 24 [12; 44] | 7% [4%; 12%] |
| All people with TB eventually cured following ACF | 611 [403; 924] | 46 [24; 84] | 8% [5%; 12%] |
| >> All people with high bacillary load TB eventually cured | 381 [252; 576] | 29 [15; 52] | 8% [4%; 12%] |
| >> All people with TB and chronic cough eventually cured | 315 [205; 483] | 18 [7; 41] | 6% [2%; 12%] |
| All people with TB cured on the first treatment attempt following ACF | 579 [381; 875] | 44 [23; 80] | 8% [5%; 12%] |
| >> All people with high bacillary load TB cured on the first treatment attempt following ACF | 352 [233; 533] | 26 [14; 48] | 8% [4%; 12%] |
| >> All people with TB and chronic cough cured on the first treatment attempt following ACF | 296 [193; 455] | 17 [6; 39] | 6% [2%; 12%] |
| All people with TB eventually cured following ACF that would have *died* in the absence of ACF | 76 [39; 132] | 6 [2; 11] | 8% [4%; 12%] |
| >> All people with high bacillary load TB eventually cured following ACF that would have *died* in the absence of ACF | 67 [32; 118] | 5 [2; 10] | 8% [4%; 12%] |
| >> All people with TB and chronic cough eventually cured following ACF that would have *died* in the absence of ACF | 43 [19; 83] | 2 [1; 7] | 6% [2%; 12%] |
| All people with TB who die due to TB | -76 [-132; -39] | -6 [-11; -2] | 8% [4%; 12%] |
| >> All people with high bacillary load TB who die due TB | -67 [-118; -32] | -5 [-10; -2] | 8% [4%; 12%] |
| >> All people with TB and chronic cough who die due TB | -43 [-83; -19] | -2 [-7; -1] | 6% [2%; 12%] |
| All people with TB that spontaneously resolve prior to receiving any treatment | -212 [-355; -126] | -17 [-33; -8] | 8% [5%; 12%] |
| >> All people with high bacillary load TB that spontaneously resolve prior to receiving any treatment | -39 [-82; -11] | -3 [-7; -1] | 8% [4%; 13%] |
| >> All people with TB and chronic cough that spontaneously resolve prior to receiving any treatment | -83 [-147; -44] | -5 [-12; -2] | 6% [2%; 12%] |

| **Population Parameter** | Benefit of ACF (using Xpert Ultra) compared to routine care [95% UR] | Additional benefit through confirmatory testing with immediate turn-around | |
| --- | --- | --- | --- |
|  |  | Absolute  [95% UR] | Relative  [95% UR] |
| Total population | 400,000 | 0 [0; 0] | 0% [0%; 0%] |
| Included in ACF efforts | 83,808 [59,388; 118,601] | 0 [0; 0] | 0% [0%; 0%] |
| All people with TB started on treatment under ACF | 870 [578; 1,294] | 69 [33; 130] | 11% [5%; 18%] |
| >> All people with high bacillary load TB started on treatment | 515 [342; 776] | 44 [21; 82] | 11% [5%; 18%] |
| >> All people with TB and chronic cough started on treatment | 473 [312; 719] | 36 [17; 68] | 11% [5%; 18%] |
| All people with TB started on treatment under ACF that would have *died* in the absence of ACF | 83 [43; 145] | 9 [4; 19] | 11% [5%; 18%] |
| >> All people with high bacillary load TB started on treatment under ACF that would have *died* in the absence of ACF | 73 [36; 130] | 8 [3; 17] | 11% [5%; 18%] |
| >> All people with TB and chronic cough started on treatment under ACF that would have *died* in the absence of ACF | 47 [21; 91] | 5 [2; 11] | 11% [5%; 18%] |
| All people with TB started on treatment under ACF that would have *spontaneously resolved* in the absence of ACF | 212 [126; 355] | 23 [10; 46] | 11% [5%; 18%] |
| >> All people with high bacillary load TB started on treatment under ACF that would have *spontaneously resolved* in the absence of ACF | 39 [11; 82] | 4 [1; 10] | 11% [5%; 18%] |
| >> All people with TB and chronic cough started on treatment under ACF that would have *spontaneously resolved* in the absence of ACF | 83 [44; 147] | 9 [4; 19] | 11% [5%; 18%] |
| All people with TB started on treatment under ACF that would have *received treatment* in the absence of ACF | 351 [227; 540] | 37 [17; 71] | 11% [5%; 18%] |
| All people with TB started on treatment under ACF that would have *received treatment, but eventually died* in the absence of ACF | 32 [20; 50] | 3 [2; 7] | 11% [5%; 18%] |
| All people with TB started on treatment under ACF that would have *received treatment and eventually be cured* in the absence of ACF | 319 [207; 491] | 34 [16; 64] | 11% [5%; 18%] |
| All people with TB eventually cured following ACF | 611 [403; 924] | 65 [31; 122] | 11% [5%; 18%] |
| >> All people with high bacillary load TB eventually cured | 381 [252; 576] | 41 [19; 76] | 11% [5%; 18%] |
| >> All people with TB and chronic cough eventually cured | 315 [205; 483] | 34 [16; 64] | 11% [5%; 18%] |
| All people with TB cured on the first treatment attempt following ACF | 579 [381; 875] | 62 [29; 116] | 11% [5%; 18%] |
| >> All people with high bacillary load TB cured on the first treatment attempt following ACF | 352 [233; 533] | 38 [18; 70] | 11% [5%; 18%] |
| >> All people with TB and chronic cough cured on the first treatment attempt following ACF | 296 [193; 455] | 32 [15; 60] | 11% [5%; 18%] |
| All people with TB eventually cured following ACF that would have *died* in the absence of ACF | 76 [39; 132] | 8 [3; 17] | 11% [5%; 18%] |
| >> All people with high bacillary load TB eventually cured following ACF that would have *died* in the absence of ACF | 67 [32; 118] | 7 [3; 15] | 11% [5%; 18%] |
| >> All people with TB and chronic cough eventually cured following ACF that would have *died* in the absence of ACF | 43 [19; 83] | 4 [2; 10] | 11% [5%; 18%] |
| All people with TB who die due to TB | -76 [-132; -39] | -8 [-17; -3] | 11% [5%; 18%] |
| >> All people with high bacillary load TB who die due TB | -67 [-118; -32] | -7 [-15; -3] | 11% [5%; 18%] |
| >> All people with TB and chronic cough who die due TB | -43 [-83; -19] | -4 [-10; -2] | 11% [5%; 18%] |
| All people with TB that spontaneously resolve prior to receiving any treatment | -212 [-355; -126] | -23 [-46; -10] | 11% [5%; 18%] |
| >> All people with high bacillary load TB that spontaneously resolve prior to receiving any treatment | -39 [-82; -11] | -4 [-10; -1] | 11% [5%; 18%] |
| >> All people with TB and chronic cough that spontaneously resolve prior to receiving any treatment | -83 [-147; -44] | -9 [-19; -4] | 11% [5%; 18%] |

| **Population Parameter** | Benefit of ACF (using Xpert Ultra) compared to routine care [95% UR] | Additional benefit through confirmatory testing with reduced test costs | |
| --- | --- | --- | --- |
|  |  | Absolute  [95% UR] | Relative  [95% UR] |
| Total population | 400,000 | 0 | 0 |
| Included in ACF efforts | 83,808 [59,388; 118,601] | 5,238 [16,271; 1,588] | 6% [2%; 22%] |
| All people with TB started on treatment under ACF | 870 [578; 1,294] | 41 [12; 129] | 6% [2%; 22%] |
| >> All people with high bacillary load TB started on treatment | 515 [342; 776] | 26 [7; 82] | 6% [2%; 22%] |
| >> All people with TB and chronic cough started on treatment | 473 [312; 719] | 21 [6; 68] | 6% [2%; 22%] |
| All people with TB started on treatment under ACF that would have *died* in the absence of ACF | 83 [43; 145] | 5 [1; 18] | 6% [2%; 22%] |
| >> All people with high bacillary load TB started on treatment under ACF that would have *died* in the absence of ACF | 73 [36; 130] | 4 [1; 16] | 6% [2%; 22%] |
| >> All people with TB and chronic cough started on treatment under ACF that would have *died* in the absence of ACF | 47 [21; 91] | 3 [1; 11] | 6% [2%; 22%] |
| All people with TB started on treatment under ACF that would have *spontaneously resolved* in the absence of ACF | 212 [126; 355] | 13 [4; 45] | 6% [2%; 22%] |
| >> All people with high bacillary load TB started on treatment under ACF that would have *spontaneously resolved* in the absence of ACF | 39 [11; 82] | 2 [0; 10] | 6% [2%; 22%] |
| >> All people with TB and chronic cough started on treatment under ACF that would have *spontaneously resolved* in the absence of ACF | 83 [44; 147] | 5 [1; 18] | 6% [2%; 22%] |
| All people with TB started on treatment under ACF that would have *received treatment* in the absence of ACF | 351 [227; 540] | 22 [6; 71] | 6% [2%; 22%] |
| All people with TB started on treatment under ACF that would have *received treatment, but eventually died* in the absence of ACF | 32 [20; 50] | 2 [1; 6] | 6% [2%; 22%] |
| All people with TB started on treatment under ACF that would have *received treatment and eventually be cured* in the absence of ACF | 319 [207; 491] | 20 [6; 64] | 6% [2%; 22%] |
| All people with TB eventually cured following ACF | 611 [403; 924] | 38 [11; 122] | 6% [2%; 22%] |
| >> All people with high bacillary load TB eventually cured | 381 [252; 576] | 24 [7; 75] | 6% [2%; 22%] |
| >> All people with TB and chronic cough eventually cured | 315 [205; 483] | 20 [6; 63] | 6% [2%; 22%] |
| All people with TB cured on the first treatment attempt following ACF | 579 [381; 875] | 36 [10; 115] | 6% [2%; 22%] |
| >> All people with high bacillary load TB cured on the first treatment attempt following ACF | 352 [233; 533] | 22 [6; 70] | 6% [2%; 22%] |
| >> All people with TB and chronic cough cured on the first treatment attempt following ACF | 296 [193; 455] | 19 [5; 59] | 6% [2%; 22%] |
| All people with TB eventually cured following ACF that would have *died* in the absence of ACF | 76 [39; 132] | 5 [1; 16] | 6% [2%; 22%] |
| >> All people with high bacillary load TB eventually cured following ACF that would have *died* in the absence of ACF | 67 [32; 118] | 4 [1; 15] | 6% [2%; 22%] |
| >> All people with TB and chronic cough eventually cured following ACF that would have *died* in the absence of ACF | 43 [19; 83] | 3 [1; 10] | 6% [2%; 22%] |
| All people with TB who die due to TB | -76 [-132; -39] | -5 [-16; -1] | 6% [2%; 22%] |
| >> All people with high bacillary load TB who die due TB | -67 [-118; -32] | -4 [-15; -1] | 6% [2%; 22%] |
| >> All people with TB and chronic cough who die due TB | -43 [-83; -19] | -3 [-10; -1] | 6% [2%; 22%] |
| All people with TB that spontaneously resolve prior to receiving any treatment | -212 [-355; -126] | -13 [-45; -4] | 6% [2%; 22%] |
| >> All people with high bacillary load TB that spontaneously resolve prior to receiving any treatment | -39 [-82; -11] | -2 [-10; 0] | 6% [2%; 22%] |
| >> All people with TB and chronic cough that spontaneously resolve prior to receiving any treatment | -83 [-147; -44] | -5 [-18; -1] | 6% [2%; 22%] |

### Figure S1 – Minimal sensitivity required for oral-swab based confirmatory testing to achieve similar epidemiological impact as sputum-based confirmatory testing.

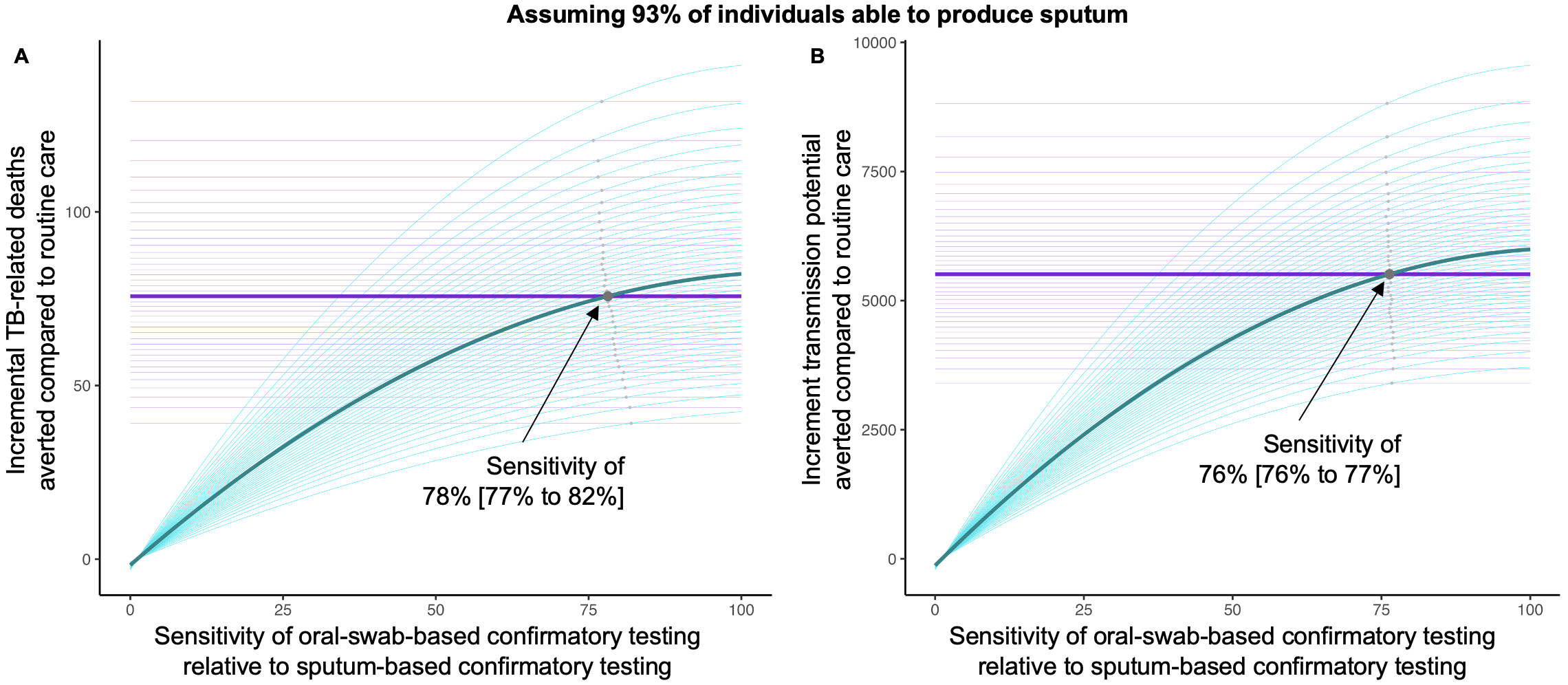

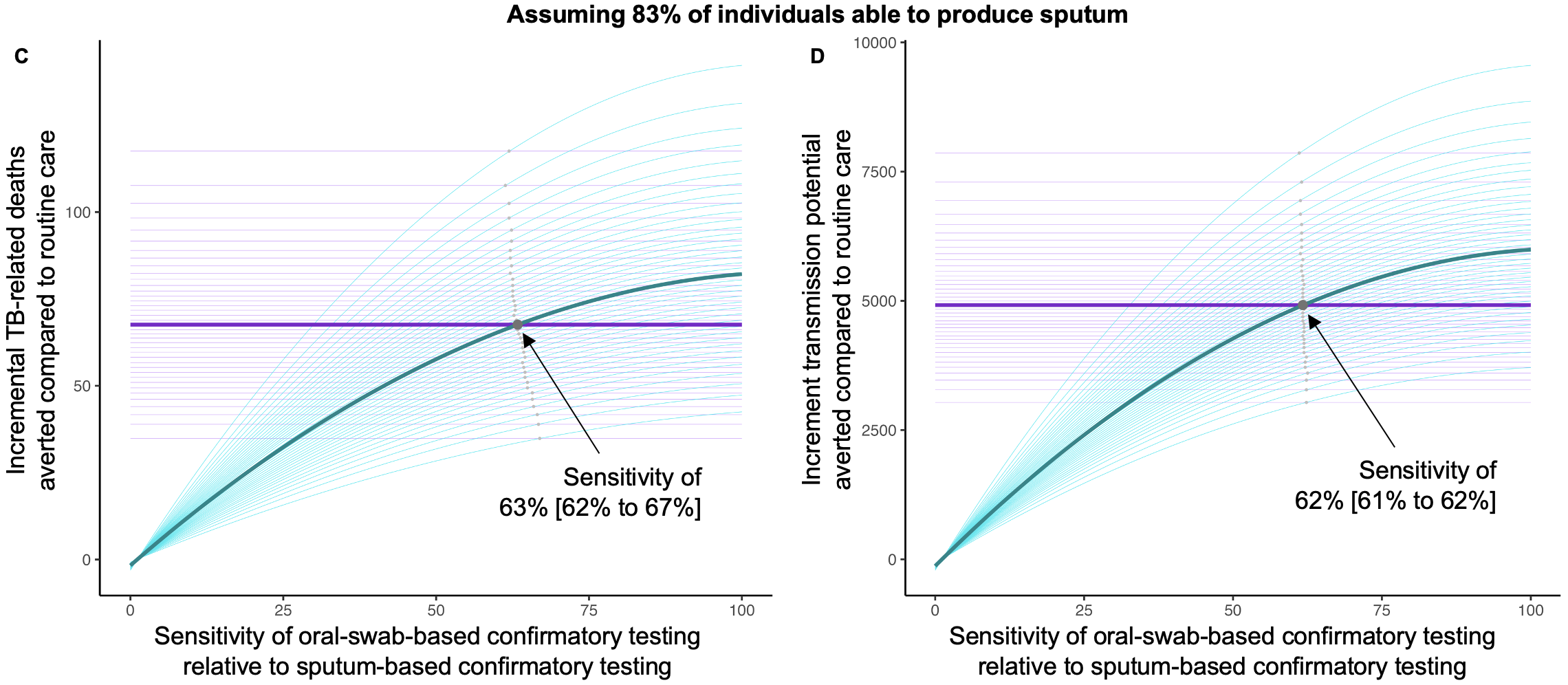

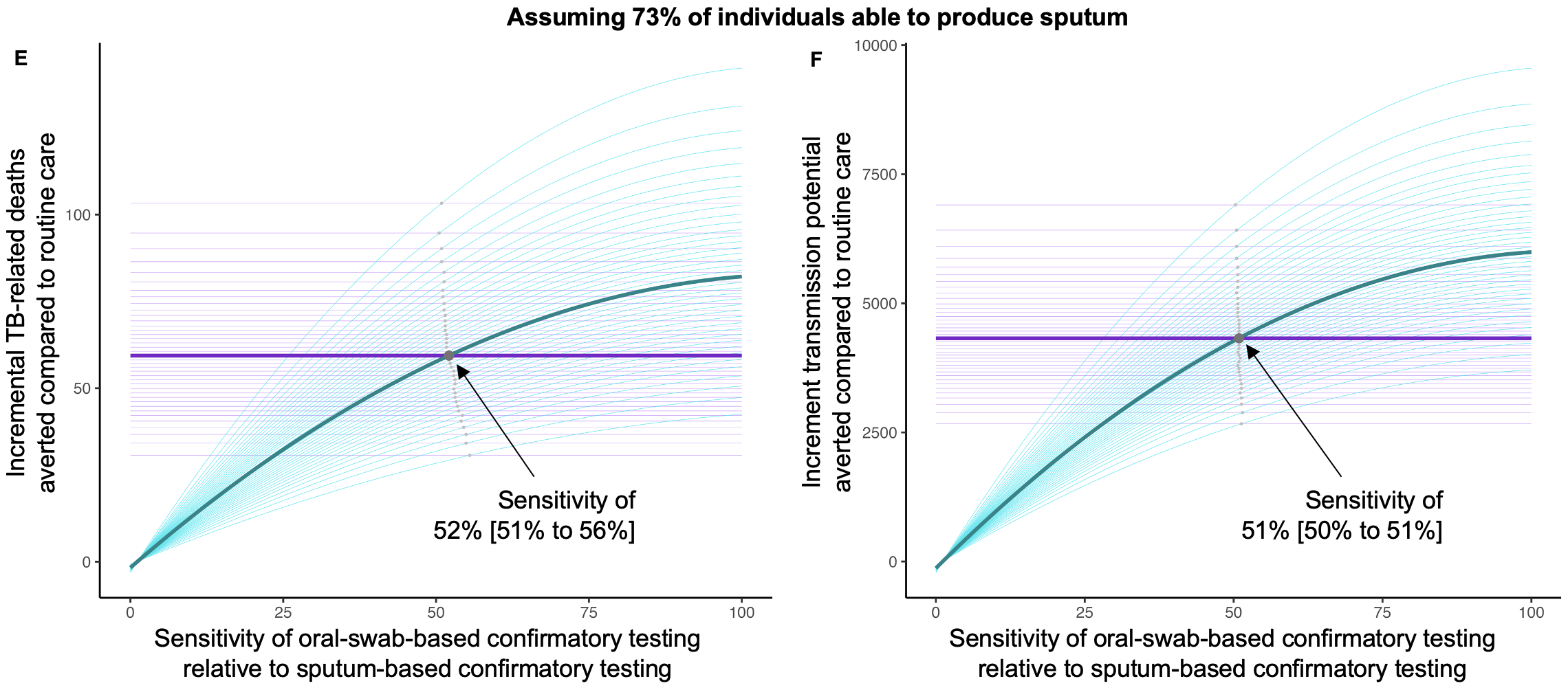

*Caption:* Evaluating the potential epidemiological impact of active case-finding (ACF) using oral-swab based confirmatory testing compared to routine care, we depict the incremental TB-related deaths averted (y-axis; panel A, C, E) and the incremental TB transmission potential averted (y-axis; panel B, D, F) at different hypothetical levels of the sensitivity of oral-swab based confirmatory testing relative to the sensitivity of sputum-based testing (x-Axis). To account for uncertainty in estimating the proportion of individuals able to produce sputum, we perform this analysis at the baseline assumption of 93% sputum production (top row of panels) as well as 83% (middle row) and 73% (bottom row) sputum production. In each panel, the thick blue line shows the median incremental number of TB deaths averted (panel A, C, E) or the median TB transmission potential averted (panel B, D, F) at different levels of sensitivity (0% to 100%) of oral-swab based confirmatory testing relative to sputum-based testing. The thick purple line represents the epidemiological benefits of ACF when using the sputum-based confirmatory test. The silver dots depict the sensitivity of oral-swab based confirmatory test, at which the epidemiological benefit of ACF using oral-swab based confirmatory testing equals the benefit of ACF using the sputum-based confirmatory test. For all panels, the thin blue and purple lines, as well as the small grey dots, respresent results within the 95% uncertainty range.

### Figure S2 – Results of the sensitivity analyses

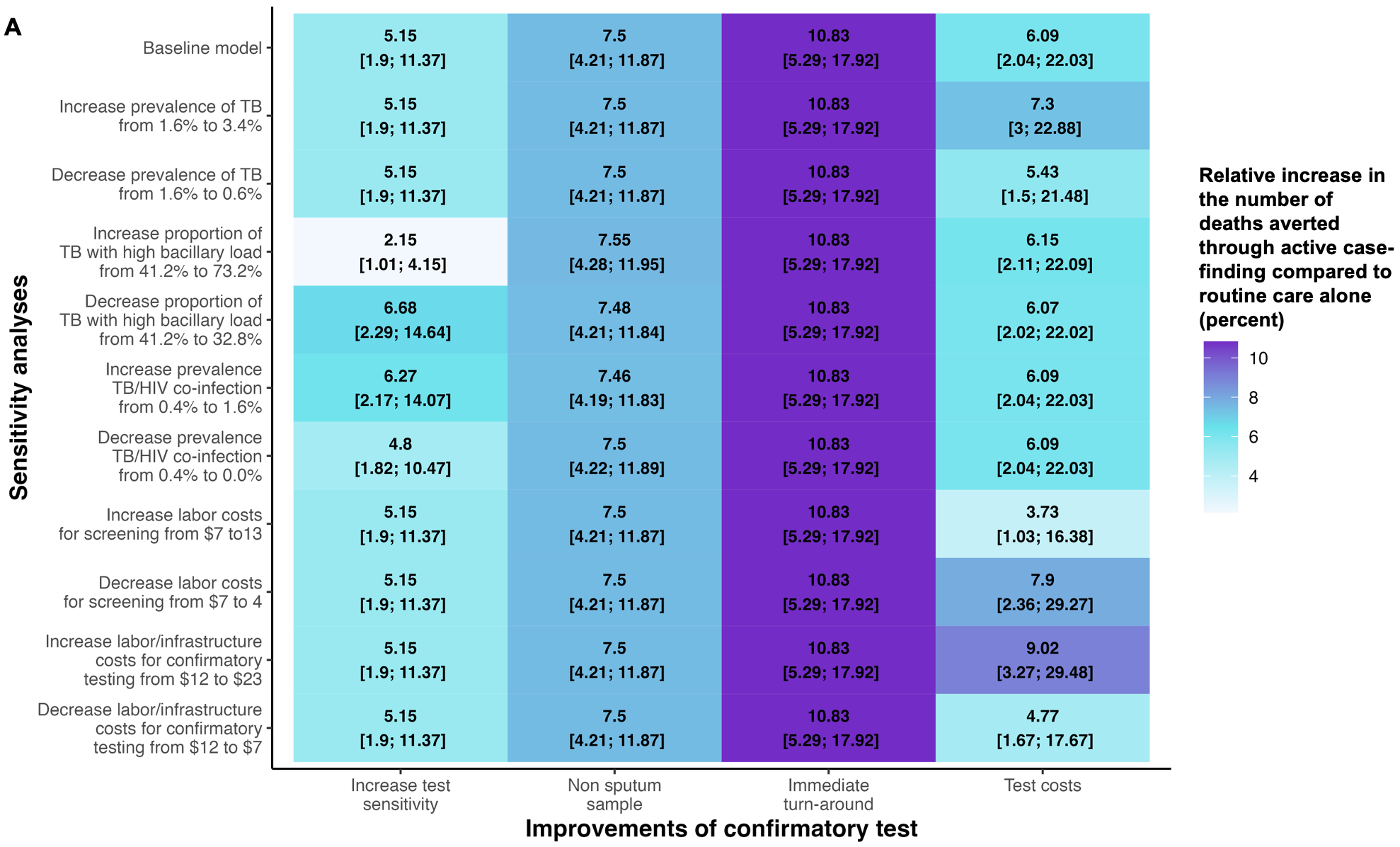

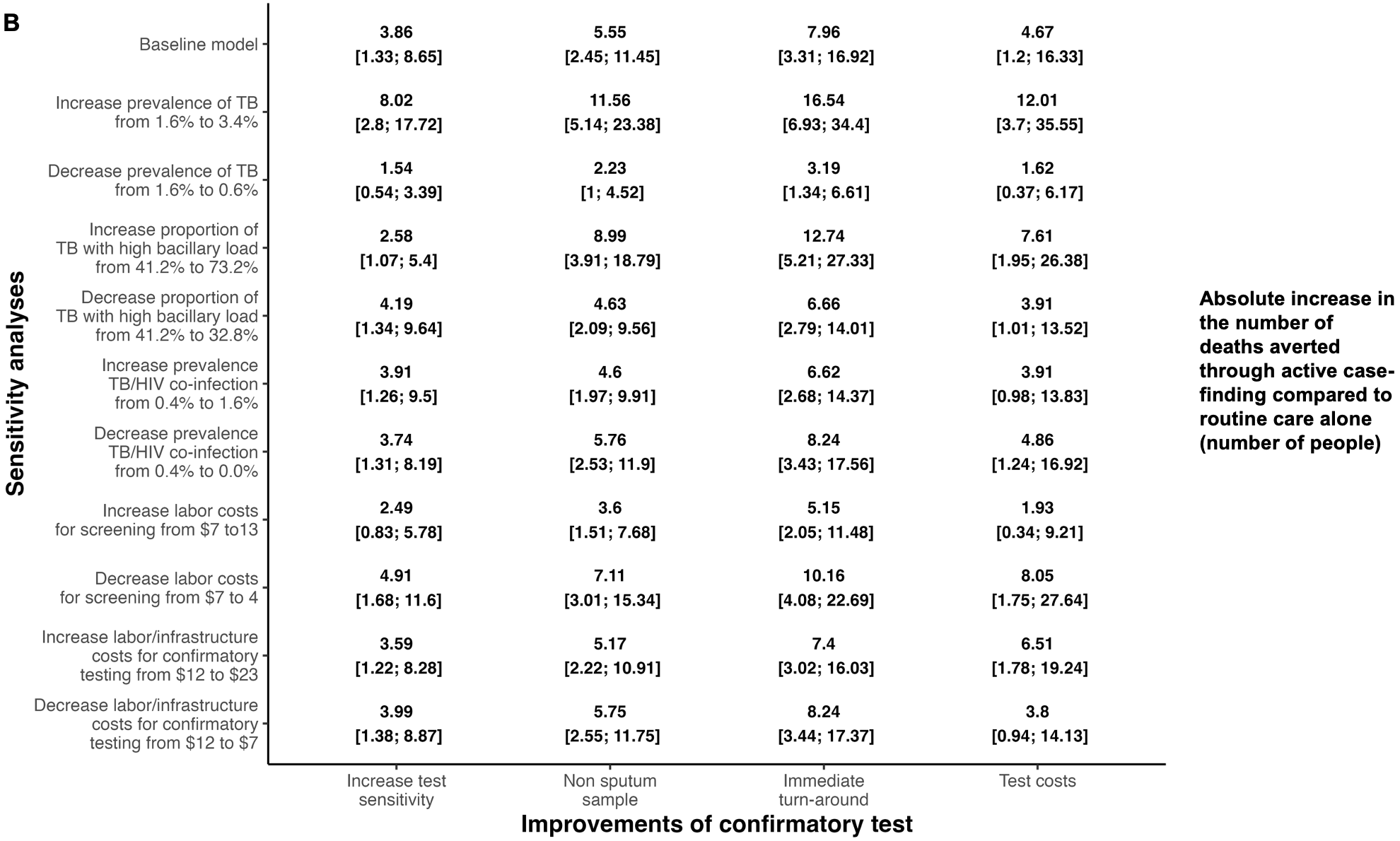

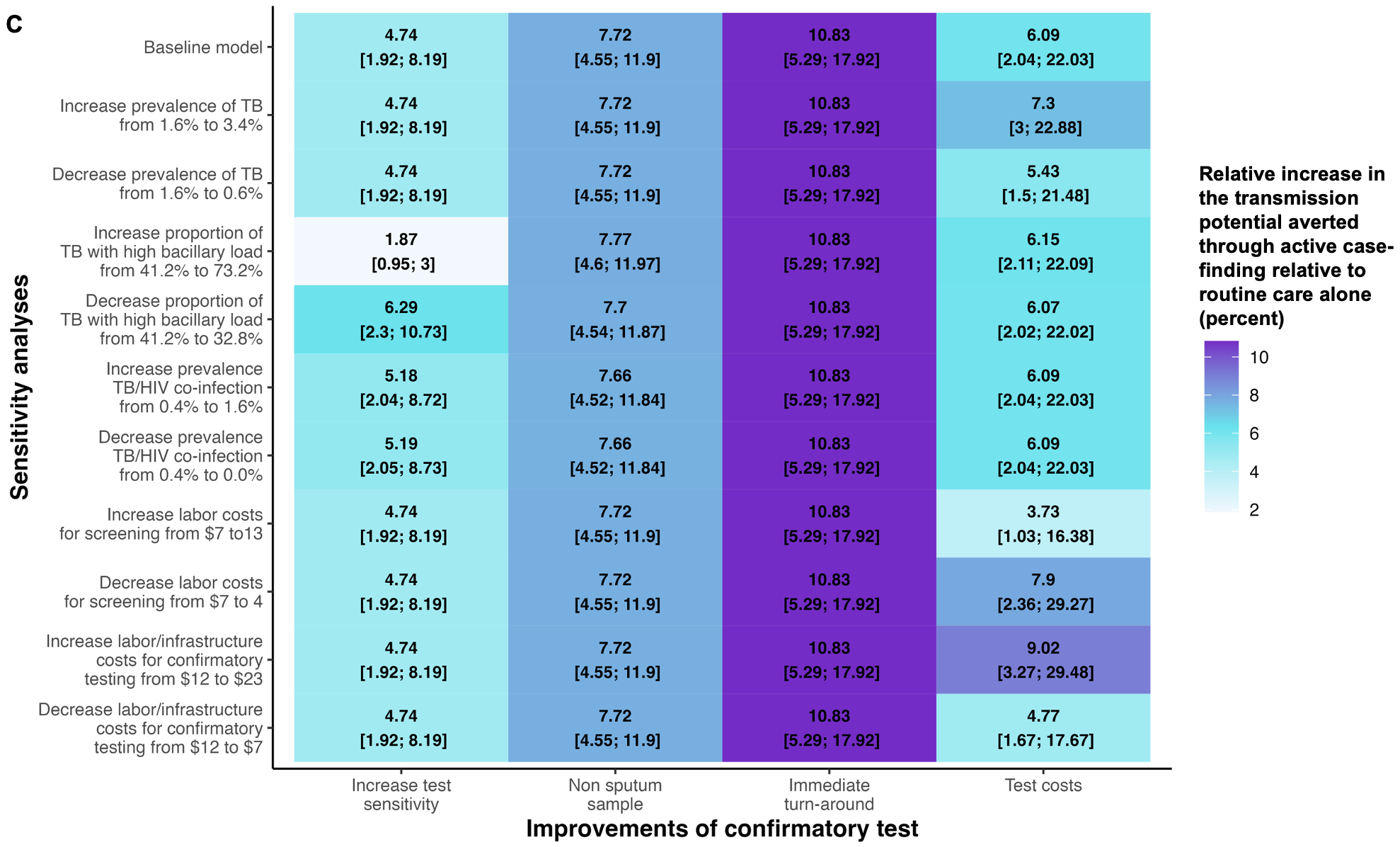

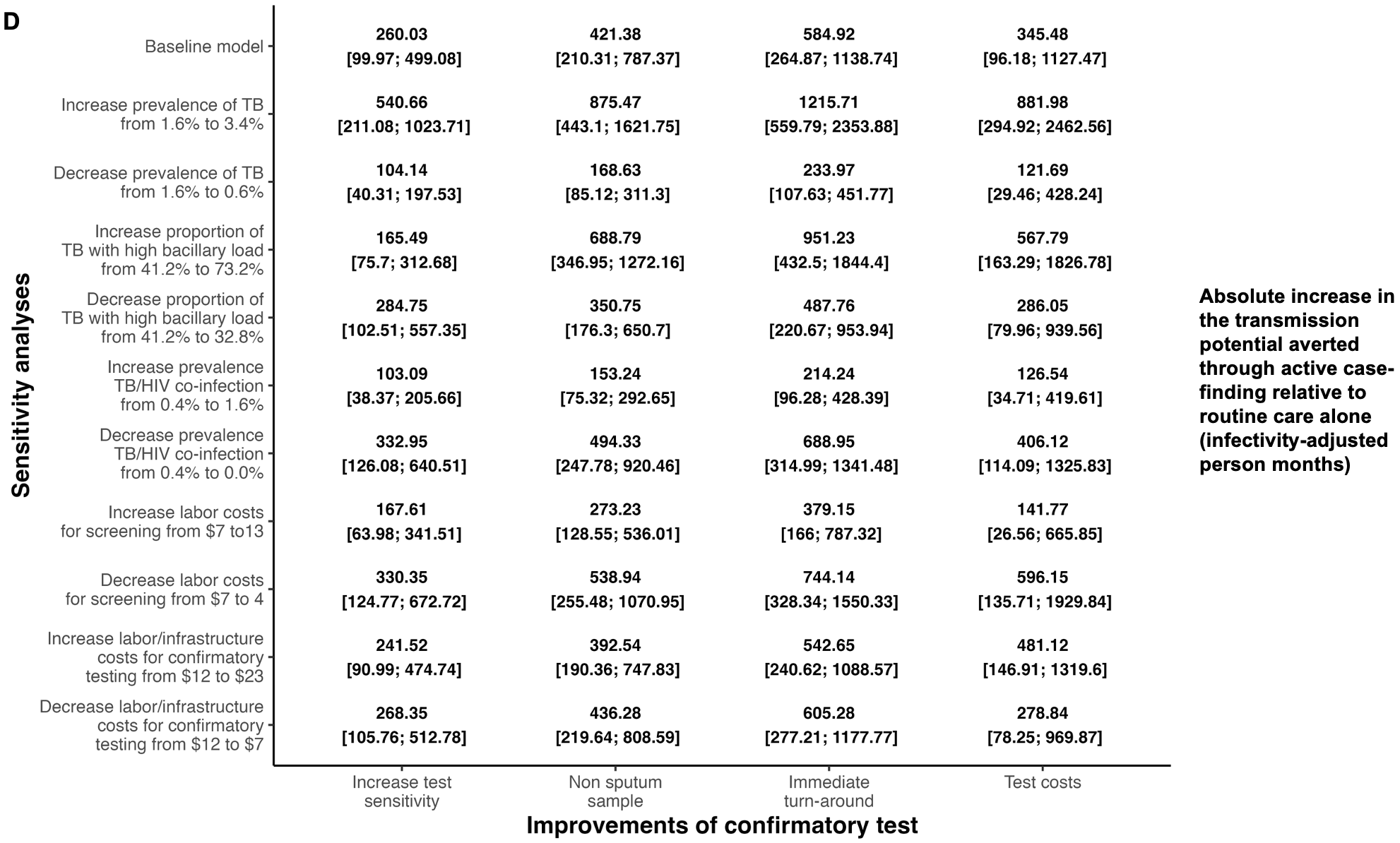

*Caption:* Shown are reductions in TB mortality (Panel A and B) and TB transmission potential (Panel C and D) through improving different characteristics of confirmatory testing (x-axis) in community-based screening, relative to active case-finding (ACF) efforts using Xpert Ultra for confirmatory testing. We present changes in these reductions when varying different exclusive model parameters (y-axis, further details in Table S5), with the median (including 95% uncertainty range) of this relative reduction denoted for each parameter-improvement combination. The term “Baseline model” refers to the impact of improving confirmatory test characteristics when keeping all parameters listed below as in the main analysis of this manuscript (Table S1 to S4 and, in the main manuscript, Table 2).

### Figure S3 - Scenario Analysis: Epidemiological effect of hypothetical improvements of a test to confirm TB during active case-finding assuming no spontaneous resolution of TB.

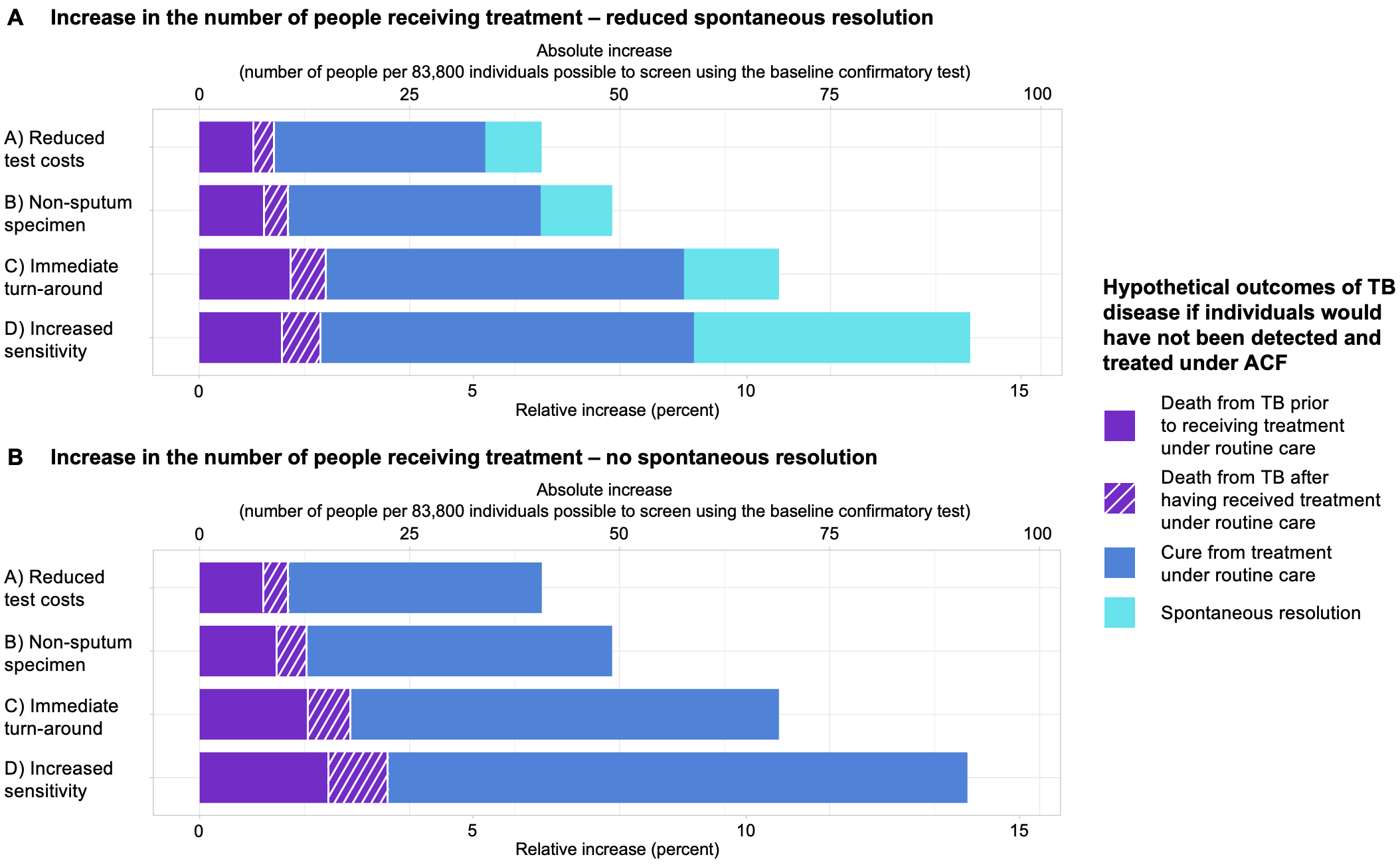

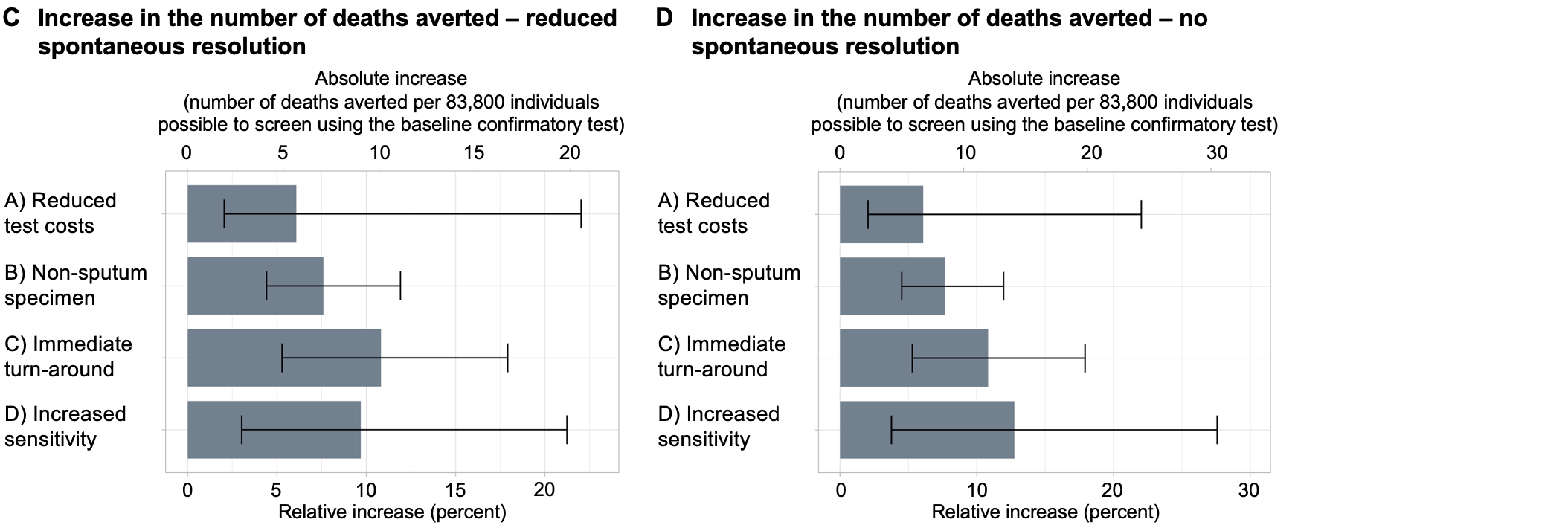

*Caption:* We depict the potential epidemiological benefit of using a hypothetically improved confirmatory test in active case-finding (ACF) efforts compared to conducting the same efforts using Xpert Ultra to confirm a positive screening test result. In contrast to the results of the baseline analysis (Figure 3, main manuscript), the results presented here assume that TB would spontaneously resolve in only half of the individuals who experience spontaneous resolution in the baseline analysis (Panel A and C), or that in no individual TB would spontaneously resolve (Panel B and D). All remaining people with TB are modeled to either be cured through treatment or to end in death (Text S5 and Table S8). Since the projections of disease durations were no longer applicable when only considering death or cure through treatment as potential outcomes of prevalent TB (Text S5), we did not evaluate the benefit of improving confirmatory test characteristics for TB transmission potential under this scenario.

### Figure S4 - Scenario Analysis: Including treatment costs as part of the active case-finding budget.

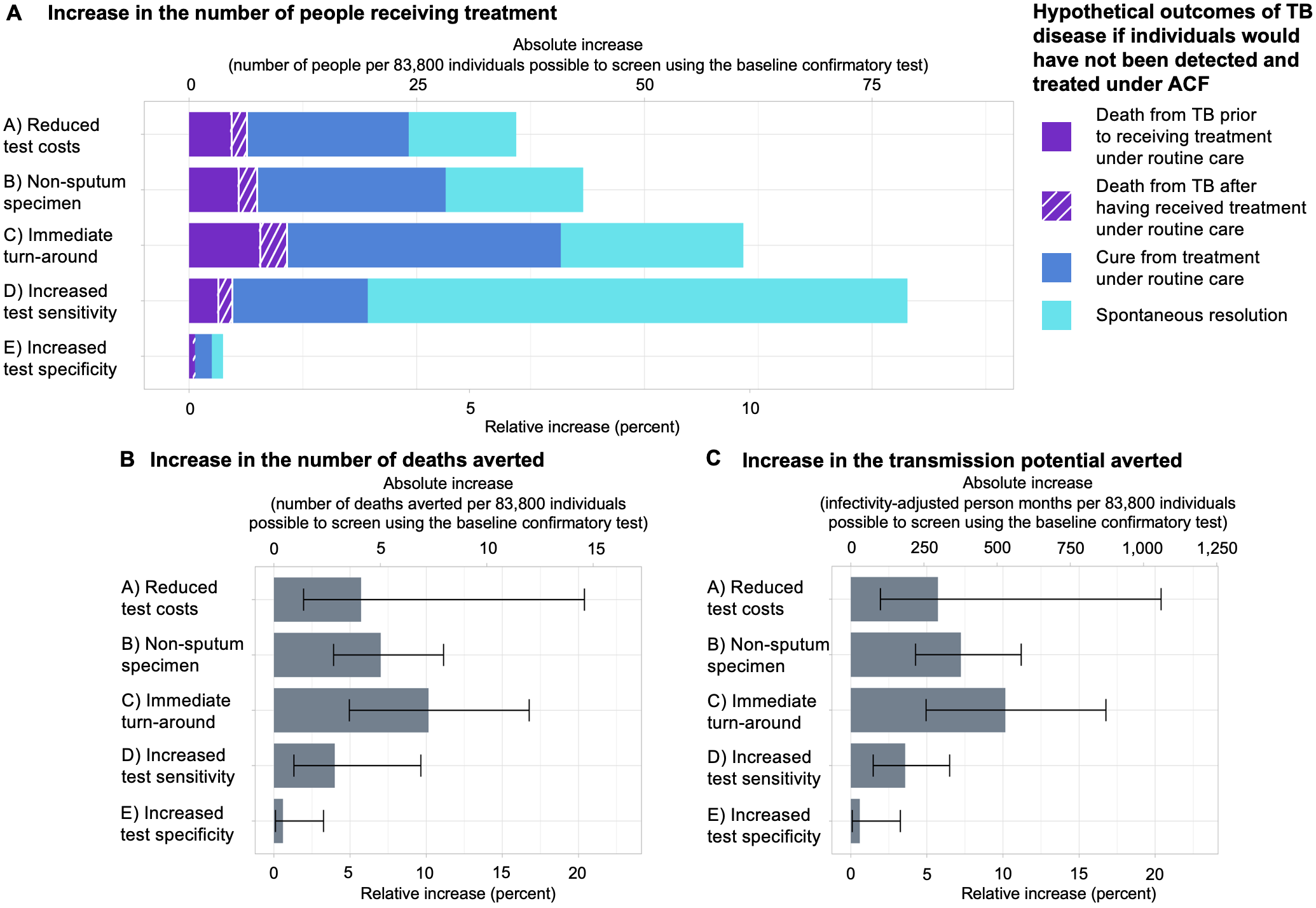

Caption: Figure S4 shows the potential epidemiological benefit of using a hypothetically improved confirmatory test in active case-finding (ACF) efforts compared to conducting the same efforts using Xpert Ultra to confirm a positive screening test result. In contrast to the results of the baseline analysis (Figure 3 in the main manuscript), the results presented here assume that treatment costs are included in the active case-finding budget, also allowing to evaluate the potential epidemiological impact of a confirmatory test with increased specificity (from 99% to 100%).

### Figure S5 - Scenario Analysis: Epidemiological effect of hypothetical improvements of a test to confirm TB during active case-finding assuming symptom screening instead of screening using chest X-ray.

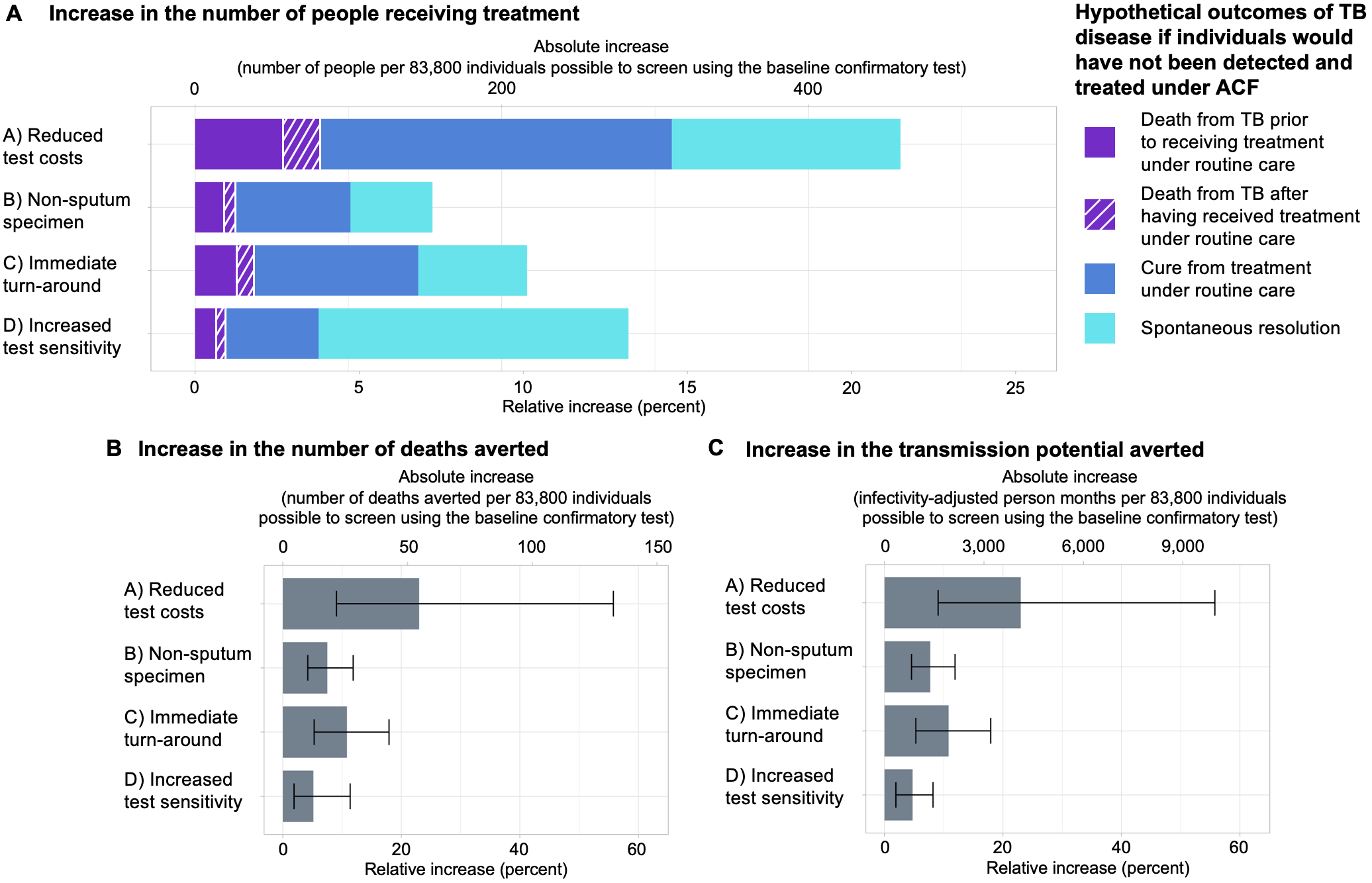

*Caption:* We present the potential epidemiological benefit of using a hypothetically improved confirmatory test in active case-finding (ACF) efforts compared to conducting the same efforts using Xpert Ultra to confirm a positive screening test result. In contrast to the results of the baseline analysis (Figure 3 in the main manuscript), the results presented here assume that symptom screening (100% sensitivity in symptomatic people, 0% in asymptomatic; 0% specificity in symptomatic people, 100% in asymptomatic; per person costs of $2.21 [$1.66-2.76]; Table S8) was conducted instead of screening using chest X-ray (overall sensitivity of 90.0%; overall specificity of 96.0%; per person costs of $10 [$7-14]; Text S1 and, in the main manuscript, Table 2).

### Figure S6 – Change in model results along the number of iterations of the monte carlo simulation performed

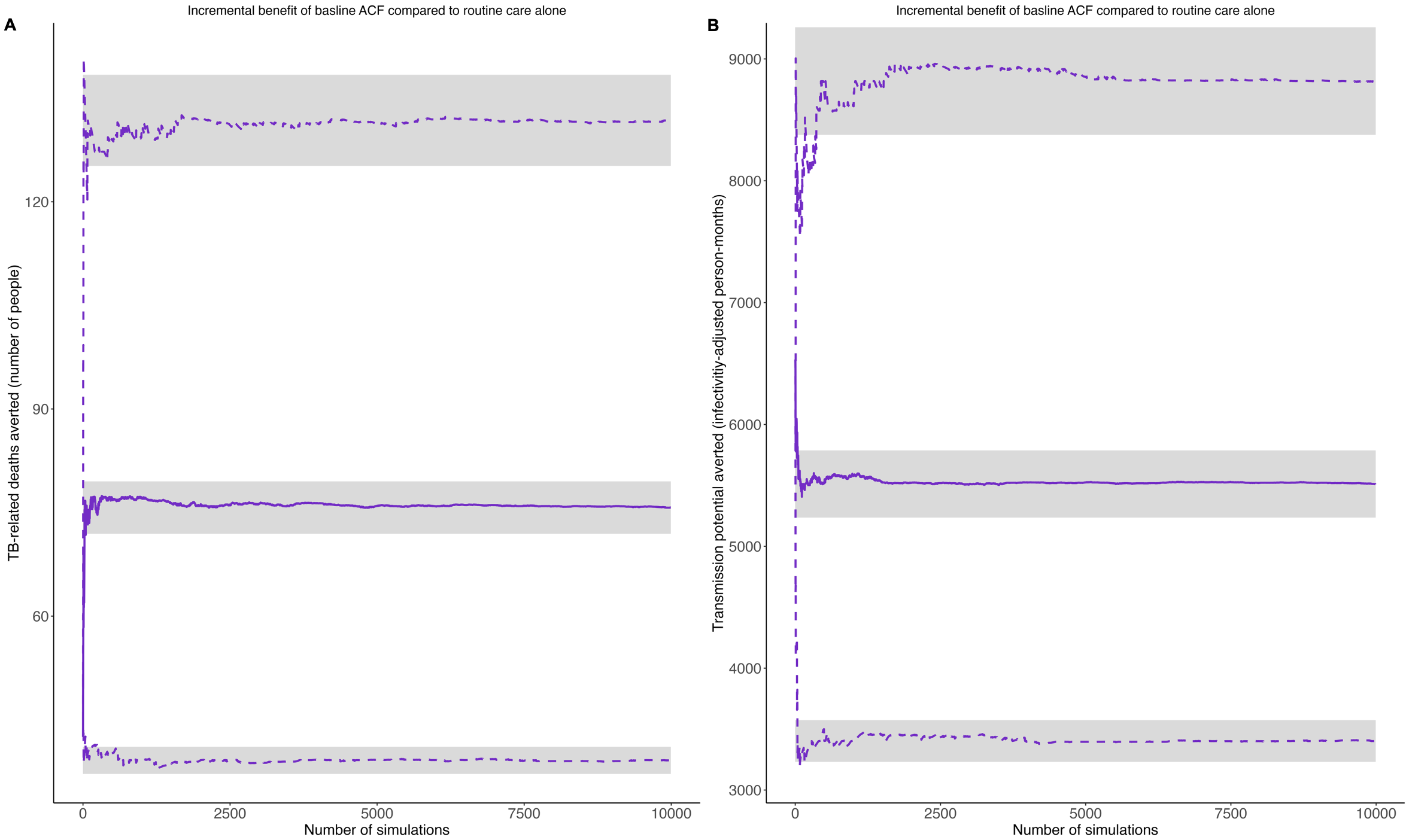

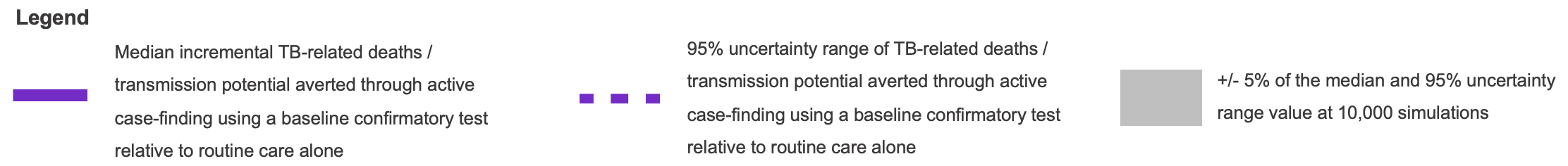

*Caption:* Shown are the changes in the median (purple line) and 2.5th (lower dashed purple line) and 97.5th (upper dashed purple line) percentiles of model results along the number of iterations of the monte carlo simulation performed. As a metric, we provide the incremental number of deaths averted (Panel A) and the incremental transmission potential averted (Panel B; in infectivity-adjusted person months) of active case-finding using a baseline confirmatory test relative to routine care alone. From less than <1,000 simulations onwards, model results stay stable within a +/- 5% range of their value at 9,992 simulations (in <0.1% of all simulations performed, random sampling led to some of the proportions of the initial population falling below zero – respective samples were dropped, resulting in 9,992 simulations being included in the final analysis).

### Sources

1.	The Republic of Uganda. The Uganda National Tuberculosis Prevalence Survey, 2014-2015 Survey Report. 2017.

2.	World Health Organization. WHO consolidated guidelines on tuberculosis. Module 2: Screening. Geneva (Switzerland); 2021.

3.	Republic of South Africa National Department of Health. The First National TB Prevalence Survey - South Africa 2018. Pretoria (South Africa); 2021.

4.	Dorman SE, Schumacher SG, Alland D, Nabeta P, Armstrong DT, King B, et al. Xpert MTB/RIF Ultra for detection of Mycobacterium tuberculosis and rifampicin resistance: a prospective multicentre diagnostic accuracy study. Lancet Infect Dis. 2018;18(1):76-84.

5.	Klinkenberg E, Floyd S, Shanaube K, Mureithi L, Gachie T, de Haas P, et al. Tuberculosis prevalence after 4 years of population-wide systematic TB symptom screening and universal testing and treatment for HIV in the HPTN 071 (PopART) community-randomised trial in Zambia and South Africa: A cross-sectional survey (TREATS). PLoS Med. 2023;20(9):e1004278.

6.	Kendall EA, Kitonsa PJ, Nalutaaya A, Erisa KC, Mukiibi J, Nakasolya O, et al. The Spectrum of Tuberculosis Disease in an Urban Ugandan Community and Its Health Facilities. Clin Infect Dis. 2021;72(12):e1035-e43.

7.	Lungu P, Kerkhoff AD, Kasapo CC, Mzyece J, Nyimbili S, Chimzizi R, et al. Tuberculosis care cascade in Zambia - identifying the gaps in order to improve outcomes: a population-based analysis. BMJ Open. 2021;11(8):e044867.

8.	Stop TB Partnership. Diagnostics, medical devices & other health products catalog 2023 [Available from: <https://www.stoptb.org/sites/default/files/gdf_diagnostics_medical_devices_other_health_products_catalog_0.pdf>.

9.	Stop TB Partnership. Artificial intelligence-powered computer-aided (CAD) software 2022 [Available from: <https://www.stoptb.org/introducing-new-tools-project/artificial-intelligence-powered-computer-aided-detection-cad-software>.

10.	Jiji Vehicels. Toyota Buses & Microbuses in Uganda 2023 [Available from: <https://jiji.ug/buses/toyota>.

11.	Global Petrol Prices. Uganda Gasoline prices, litre, 21-Aug-2023 2023 [Available from: <https://www.globalpetrolprices.com/Uganda/gasoline_prices/#:~:text=Uganda%3A%20The%20price%20of%20octane,see%20the%20prices%20in%20gallons>.

12.	Baik Y, Nakasolya O, Isooba D, Mukiibi J, Kitonsa PJ, Erisa KC, et al. Cost to perform door-to-door universal sputum screening for TB in a high-burden community. Int J Tuberc Lung Dis. 2023;27(3):195-201.

13.	The Global Fund. Global Fund, Stop TB Partnership and USAID Announce New Collaboration with Danaher to Reduce Price and Increase Access to Cepheid’s TB Test. 2023.

14.	Thompson RR, Nalugwa T, Oyuku D, Tucker A, Nantale M, Nakaweesa A, et al. Multicomponent strategy with decentralised molecular testing for tuberculosis in Uganda: a cost and cost-effectiveness analysis. Lancet Glob Health. 2023;11(2):e278-e86.

15.	Kairu A, Orangi S, Oyando R, Kabia E, Nguhiu P, Ong Ang OJ, et al. Cost of TB services in healthcare facilities in Kenya (No 3). Int J Tuberc Lung Dis. 2021;25(12):1028-34.

16.	World Bank. GDP deflator (base year varies by country) - United States 2023 2023 [Available from: <https://data.worldbank.org/indicator/NY.GDP.DEFL.KD.ZG?locations=US>.

17.	Ryckman TS, Dowdy DW, Kendall EA. Infectious and clinical tuberculosis trajectories: Bayesian modeling with case finding implications. Proc Natl Acad Sci U S A. 2022;119(52):e2211045119.

18.	Ku CC, MacPherson P, Khundi M, Nzawa Soko RH, Feasey HRA, Nliwasa M, et al. Durations of asymptomatic, symptomatic, and care-seeking phases of tuberculosis disease with a Bayesian analysis of prevalence survey and notification data. BMC Med. 2021;19(1):298.

19.	World Health Organization. Tuberculosis data, CVS files to download Geneva (Switzerland)2023 [Available from: <https://www.who.int/teams/global-tuberculosis-programme/data>.

20.	Kendall EA, Shrestha S, Cohen T, Nuermberger E, Dooley KE, Gonzalez-Angulo L, et al. Priority-Setting for Novel Drug Regimens to Treat Tuberculosis: An Epidemiologic Model. PLoS Med. 2017;14(1):e1002202.

21.	Knight GM, Gomez GB, Dodd PJ, Dowdy D, Zwerling A, Wells WA, et al. The Impact and Cost-Effectiveness of a Four-Month Regimen for First-Line Treatment of Active Tuberculosis in South Africa. PLoS One. 2015;10(12):e0145796.

22.	Velayutham B, Chadha VK, Singla N, Narang P, Gangadhar Rao V, Nair S, et al. Recurrence of tuberculosis among newly diagnosed sputum positive pulmonary tuberculosis patients treated under the Revised National Tuberculosis Control Programme, India: A multi-centric prospective study. PLoS One. 2018;13(7):e0200150.

23.	United Nations. Crude death rate per country. Manhattan (New York, United States); 2023.

24.	UNAIDS. AIDSinfo – Global data on HIV epidemic and response Geneva (Switzerland)2023 [Available from: <https://aidsinfo.unaids.org>.

25.	Akessa GM, Tadesse M, Abeb G. Survival Analysis of Loss to Follow-Up Treatment among Tuberculosis Patients at Jimma University Specialized Hospital, Jimma, Southwest Ethiopia. International Journal of Statistical Mechanics. 2015;2015.

26.	Fox GJ, Nguyen VN, Dinh NS, Nghiem LPH, Le TNA, Nguyen TA, et al. Post-treatment Mortality Among Patients With Tuberculosis: A Prospective Cohort Study of 10 964 Patients in Vietnam. Clin Infect Dis. 2019;68(8):1359-66.

27.	Andama A, Whitman GR, Crowder R, Reza TF, Jaganath D, Mulondo J, et al. Accuracy of Tongue Swab Testing Using Xpert MTB-RIF Ultra for Tuberculosis Diagnosis. J Clin Microbiol. 2022;60(7):e0042122.

28.	Steadman A, Andama A, Ball A, Mukwatamundu J, Khimani K, Mochizuki T, et al. New manual qPCR assay validated on tongue swabs collected and processed in Uganda shows sensitivity that rivals sputum-based molecular TB diagnostics. Clin Infect Dis. 2024.

29.	Federal Republic of Nigeria. First National TB Prevalence Survey 2012, Nigeria. Abuja, Nigeria; 2012.

30.	Pooran A, Theron G, Zijenah L, Chanda D, Clowes P, Mwenge L, et al. Point of care Xpert MTB/RIF versus smear microscopy for tuberculosis diagnosis in southern African primary care clinics: a multicentre economic evaluation. Lancet Glob Health. 2019;7(6):e798-e807.

31.	Ministry of Health & Family Welfare - Government of India. India Tuberculosis Report. 2022.

32.	Cambodia Ministry of Health. Report of the second national tuberculosis prevalence survey, 2011. Phnom Penh; 2012.

33.	Sekandi JN, List J, Luzze H, Yin XP, Dobbin K, Corso PS, et al. Yield of undetected tuberculosis and human immunodeficiency virus coinfection from active case finding in urban Uganda. Int J Tuberc Lung Dis. 2014;18(1):13-9.

34.	Muniyandi M, Lavanya J, Karikalan N, Saravanan B, Senthil S, Selvaraju S, et al. Estimating TB diagnostic costs incurred under the National Tuberculosis Elimination Programme: a costing study from Tamil Nadu, South India. Int Health. 2021;13(6):536-44.

35.	Machekera SM, Wilkinson E, Hinderaker SG, Mabhala M, Zishiri C, Ncube RT, et al. A comparison of the yield and relative cost of active tuberculosis case-finding algorithms in Zimbabwe. Public Health Action. 2019;9(2):63-8.

36.	The Republic of Uganda. Uganda population-based HIV impact assessment. 2022.

37.	World Health Organization. Global Tuberculosis Report 2014. Geneva; 2014.

38.	National AIDS Control Organization & ICMR-National Institute of Medical Statistics. India HIV Estimates 2021: Fact Sheet. New Delhi, India: Ministry of Health and Family Welfare, Government of India; 2022.
